# HPV binding antibodies are associated with protection: a meta-analysis

**DOI:** 10.1101/2025.11.26.25341002

**Authors:** Matthew T Berry, Matthew R Palmer, Shanchita R Khan, Ellen Bowden-Reid, Ece Egilmezer, Megan A Smith, Deborah Bateson, Karen Canfell, Miles P Davenport, David S Khoury

**Affiliations:** Kirby Institute, University of New South Wales, Sydney, New South Wales, Australia; Cancer Elimination Collaboration, Sydney School of Public Health, University of Sydney, Sydney, New South Wales, Australia; Sydney Medical School, University of Sydney, Sydney, New South Wales, Australia

**Keywords:** Correlates of Protection, Human Papillomavirus, Vaccine Efficacy, Antibodies

## Abstract

**Background:** Human papillomavirus (HPV) is the major cause of cervical cancer globally. Current vaccines have shown high efficacy at preventing HPV infection and other surrogate disease endpoints in randomised controlled trials (RCT). However, uptake is limited by accessibility and cost. Rapid clinical evaluation is critical for shortening time-to-market and encouraging the development and deployment of affordable vaccines. We aim to establish a validated correlate of protection that will support vaccine developers and decision makers with vaccine development, approval, dose schedules and deployment decisions.

**Methods:** We utilised three existing Cochrane Systematic Reviews of RCTs of HPV vaccines and extended these to 1^st^ January 2026. We identified and extracted data from trials where both efficacy and immunogenicity (HPV-specific antibodies) were reported. For HPV types where vaccine efficacy was reported but not immunogenicity (Types 31/33/45/52/58) we sourced antibody binding concentrations from separate immunogenicity studies. We linked antibody binding concentrations from each RCT to the corresponding vaccine efficacy data, matching by study, vaccine arm, number of doses, and HPV type. We use a Bayesian framework in a model-based meta-analysis to examine the association between antibody binding concentrations and vaccine efficacy.

**Findings:** Combining data on both vaccine-targeted and non-targeted HPV types, we find an association between type-specific antibody binding concentrations and vaccine efficacy against the three HPV outcomes tested: incident infection, persistent infection and grade 2 or 3 cervical intraepithelial neoplasia (CIN2+) (*p* < 0.0005, for all outcomes). We also compared this association with data from studies of natural infection, which had shown an association between antibody binding concentrations and risk of infection. This demonstrated that the association between vaccine-induced antibodies and vaccine efficacy was well aligned with the existing HPV literature on correlates of risk from natural exposure. Finally, we find evidence that the antibody concentrations associated with 50% (of the maximal) vaccine efficacy differ by type (*p* < 0.0005, for example 16 sIU/ml for HPV58, but < 0.5 IU/ml for HPV 16/18).

**Interpretations:** HPV type-specific antibody binding concentrations are associated with protection against HPV related clinical outcomes, across different HPV types and vaccines, and after natural infection. The consistent observation that antibody binding concentrations correlate with protection provides support for the use of antibody binding for immunobridging, for example in non-inferiority studies when assessing similar vaccines.

**Funding:** This work is supported by National Health and Medical Research Council of Australia and the University of New South Wales.

## Introduction

*Human papillomavirus* (HPV) is the major cause of cervical cancers globally^1,2^. There are many different types of HPV, of which 8 types contribute to a significant proportion of cancer cases^3,4^. The disease burden is not evenly distributed across these oncogenic HPV types, with HPV16 and HPV18 being responsible for the greatest number of cervical cancers globally (60% and 15%, respectively)^5^. Vaccines targeting two or more HPV types have all been developed and widely used^6–8^ – have demonstrated to be highly effective at reducing the incidence of HPV infection, persistent HPV infection, and grades 2 and 3 cervical intraepithelial neoplasia (CIN2/3) with some results indicating that vaccines have reduced rates of cervical cancers^9–12^.

At the time of writing, there are six licensed HPV vaccines^13^. There is considerable demand for these vaccines and vaccine availability can limit uptake, especially in low- and middle-income countries (LMIC). The development and deployment of affordable vaccines could improve access. However, given the availability of effective vaccines, there are many practical and ethical barriers to conducting placebo-controlled trials to assess the efficacy of new vaccines or regimens. Further, clinical non-inferiority trials require very large trials. These challenges would be reduced by the establishment of a validated immune correlate of protection for HPV vaccination^14^. An immune correlate of protection is an immune biomarker (typically serological) that has been shown to correlate with vaccine effect. This may be a specific immune threshold above which individuals are thought to be protected (threshold of protection)^15^, or more commonly is a continuous association between an immune marker and protection^16,17^. Demonstrating a robust correlate of protection would support for example the use of immunogenicity-based non-inferiority endpoints in clinical trials as a path to vaccine assessment and licensure. In addition, a correlate of protection for HPV could also support public health decision makers.

In this study we aimed to identify if there is a relationship between HPV-specific antibody concentrations after vaccination and vaccine efficacy that may function as a correlate of protection. Leveraging existing systematic reviews of vaccine efficacy, we aggregated data from all available randomised controlled trials (RCTs) of vaccine efficacy where antibody responses were also assessed. In a model-based meta-analysis, we examined the relationship between antibody concentrations and protection against HPV-related outcomes in RCTs. We test the robustness and generalisability of this association against independent data on antibody responses and protection after natural infection. Finally, we explore the robustness of this relationship across HPV types.

## Methods

### Systematic search

To conduct our systematic search of the literature, we used three existing Cochrane Systematic Reviews^18–20^ as the basis to begin our search. From these three reviews we screened all articles included in the reviews and also those articles identified by the reviews but listed as excluded in the analysis. In addition to these reviews, we extend the systematic search for more recent articles. Using the search terms stated in the supplementary material (Table S1) we searched PubMed and Embase. Our search consisted of all results between 7 January 2022 and 1 January 2026.

Each article was screened independently by two authors (from a team of four authors: MTB, MRP, EBR and EE) using the following inclusion criteria (extended details in supplementary methods Table S2):

- Articles reported data from a randomised controlled trial (RCT)
- RCT needed to use an HPV vaccine with control group. Controls could be either placebo or an irrelevant vaccine.
- RCT needed to measure immunogenicity (measure of antibodies).
- RCT needed to report vaccine efficacy against an HPV-related outcome.
- Articles were excluded if they did not provide either immunogenicity or efficacy data.

### Data extraction

Data extraction was performed by four authors (MTB, MRP, SRK or EE). One author would extract the data, which was then checked for accuracy by a different author. For vaccine efficacy data, the case count data, disaggregated by the smallest reported time intervals, were extracted against any outcome that was reported in the article. For each RCT there were often multiple articles and reports of case numbers in the trial, but only the most temporally disaggregated data for a given outcome from an RCT was used, to avoid using duplicated data (Table S3). Immunogenicity data was extracted by one reviewer and then checked by a second reviewer. In some instances, immunogenicity data was reported in multiple papers relating to the one RCT. In these cases, all available immunogenicity time point data were used, but where immunogenicity data at a particular timepoint was reported in two or more studies, only the immunogenicity data reported in one of the papers was used (i.e. the paper containing the most immunogenicity time points was selected in such cases). All immunogenicity data was extracted in the units provided in the respective papers.

### Conversion of antibody binding concentrations to international units

Where the extracted antibody binding data was not reported in international units, antibody binding concentrations were converted to IU/ml. The conversion factors for converting antibody binding concentrations from the units used in a given assay to international units were obtained from the literature (Table S4)^21,22^ (ensuring the assays are the same and performed by the same authors). Where a conversion factor for an assay could not be identified in the literature these were not included in the primary analysis.

For HPV types 6, 11, 31, 33, 45, 52 and 57, we did not have results in international units (these standards are currently being developed^23^). To include these HPV types in our analysis we introduce the concept of scaled international units (sIU/ml). For HPV types 16 and 18, the scaled international units are simply the international units (i.e. unchanged). For all other types, we calculate the average fold-difference, for each vaccine regimen, for the corresponding HPV type compared to HPV16 (details in supplement). These average fold-differences are applied to the binding antibody concentration to HPV16 (in international units), to obtain the scaled international units for each type.

### Meta-analysis of vaccine effect

In our meta-analysis we use the reported number of HPV related events extracted from each study (in the vaccine arm/s and control arm) to estimate an aggregate vaccine effect for each vaccine regimen against each HPV type. To do this, we used a Bayesian meta-regression approach, assuming a binomial distribution in the number of cases in each arm (with a unique parameter capturing the probability of cases in the control arm for each study), and assuming a common vaccine effect (relative risk, RR) for each vaccine regimen against each HPV type (across all studies with the same vaccine regimen).

### Model-based meta-analysis for a correlate of protection

When fitting a relationship between vaccine effect and antibody concentrations, we used a similar approach to that described above, but instead of estimating an aggregate relative risk parameter for each vaccine regimen, we assume the relative risk (i.e. vaccine effect) was described by a three-parameter logistic model of the antibody binding concentrations. This is similar to the model-based meta-analysis approach we have used previously^16,17,24^. Specifically, we employ the maximal logistic function,

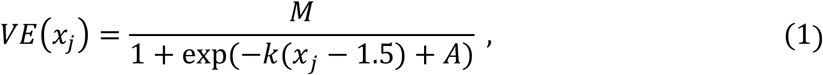

where *x*_j_, is the logarithm (base 10) of the type-specific antibody concentration for HPV type *j*, *k* is the slope of the logistic function, *M*, is the maximum vaccine efficacy (i.e. 1-RR), *A*, infers the *EC*_50_ value (i.e. the antibody concentration associated with half the maximal vaccine effect). This equation relates the antibody concentration to protection, enabling a plateau where high protection is observed for a range of antibodies. We applied our Bayesian approach for modelling the relationship between antibody binding concentrations and vaccine efficacy using all data across different vaccines, regimens, timepoints, and HPV types (supplementary methods).

To expand the model to consider type-specific correlates of protection we expanded this model to include a unique parameter *A*_ℎ_ for each HPV type, i.e.

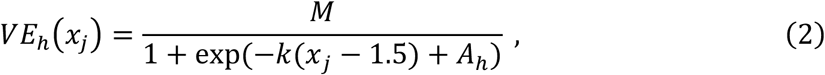

where, *VE*_ℎ_ is the vaccine efficacy against HPV type ℎ, with the parameter *A*_ℎ_ allowing the *EC*_50_ for each HPV type ℎ to differ. Under this model, the slope, *k*, and maximum efficacy *M*, are the same across all HPV types. In addition, in a sensitivity analysis, we included an effect for assay in our analysis by replacing the *A* (equation 1) with *A*_w_ and allowed the model to estimate a unique *A*_w_ for each assay. This is equivalent to estimating an assay specific *EC*_50_.

### Bayesian model fitting and statistical analysis

Model priors were selected as described in the supplementary methods (section S4). Posterior samples were computed for all model parameters using RStan (version 2.32.2)^25,26^ and the default HMC sampler. Prior predictive checks were used to assess the impact of prior choice on posterior estimates. All meta-analysis results are reported as the median of the posterior along with the central 95% credible interval. To determine if this correlation is significant, we conduct a test analogous to a one-sided hypothesis test that the slope parameter is less than or equal to 0, i.e. *p* = *P*(*k* ≤ 0) (a slope equal to zero would have no correlation). In this case we reject the null hypothesis if *p* is below our threshold (*α* = 0.05, i.e. when *p* < *α*). All analysis was conducted using R version 4.4.1.^27^

### Ethics Declarations

This work was approved under the UNSW Sydney Human Research Ethics Committee (approval HC200242).

## Results

### Vaccine efficacy and immunogenicity over time

Using the results of three systematic reviews of HPV vaccine efficacy ^18–20^ and our expanded search (7^th^ January 2022 to 1^st^ January 2026), we identified 17 RCTs (and 87 publications relating to these 17 trials) that met the inclusion criteria (Figure S1, Table S5). Two of these trials were excluded from our meta-analysis as they included only men and thus differed considerably from the 15 other studies. Within the 15 trials, 13 trials either used an antibody assay where a conversion to international units was possible or reported immunogenicity in international units. Thus, we excluded two trials from our analysis of correlates of protection as antibody binding concentrations were in non-comparable units (one trial was of the monovalent vaccine^28^ and one trial used the Cecolin vaccine^29^). Within the remaining 13 trials, all but one reported the efficacy of a single vaccine (i.e. Cervarix (bivalent) (n=6), Gardasil (quadrivalent) (n=6)), and one study reported the efficacy of two different vaccines (Cervarix (bivalent) and Gardasil-9 (nonavalent)). A range of clinical outcomes were reported with the most commonly reported outcomes being: incident cervical HPV infection (n=6), persistent HPV infection for 6 months (n=11), and CIN2+ (n=9) (Table S6, Figure S2-S9).

To test for an association between antibody binding concentration and vaccine efficacy, we aggregated the immunogenicity and efficacy data across all the available RCTs, matching efficacy and immunogenicity by vaccine schedule and time points. Initially, we only consider HPV 16 and 18, where we have defined international units. We chose to focus on the vaccine efficacy estimates against incident HPV infection, persistent HPV infection (6 months) and CIN2+, since these were the most commonly reported outcomes. Vaccine efficacy against all three HPV-related outcomes (and against the HPV types targeted by the vaccines) was high across three dose, two dose and one dose regimens and remained high out to 10 years post-vaccination where this was reported (Figure 1A, Figure S10). This high efficacy was maintained over time despite declining antibody concentrations (Figure 1B). Matching the geometric mean antibody binding concentrations (GMCs) data from each RCT to the corresponding vaccine efficacy data, by timepoint (since vaccination) and HPV type, we see that vaccine efficacy does not increase with antibody titres for any of the three outcomes tested (Figure 1C, Figure S11, e.g. efficacy appears to decrease with antibody concentration for persistent infection). However, given that all efficacy estimates were high across vaccines, number of doses, time and HPV types, finding a correlate of protection was not likely with this data. Instead, at least some situation or group where lower vaccine efficacy is observed is needed in order to explore correlates of protection.

**Figure 1:**
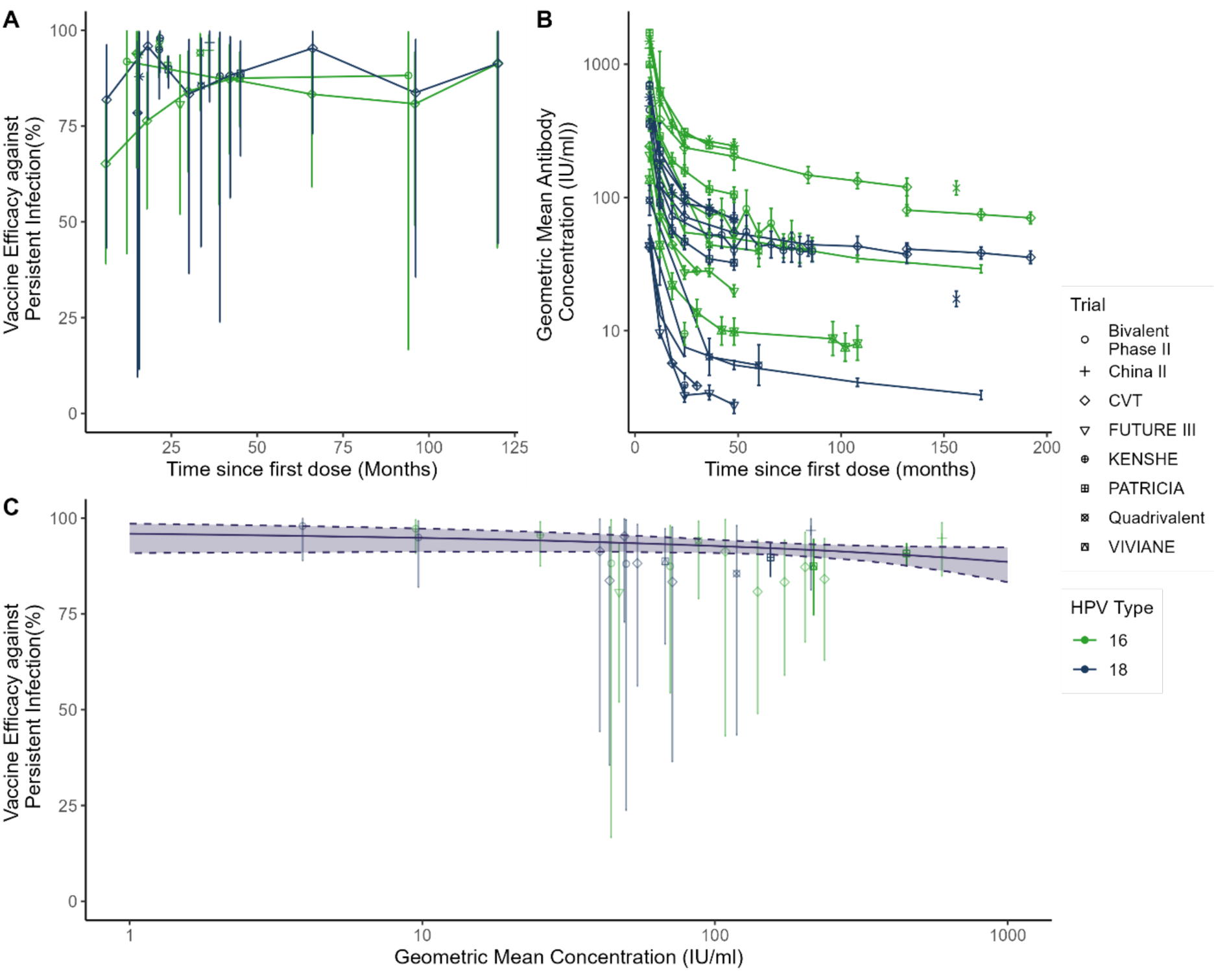
HPV vaccine efficacy and immunogenicity. A) Estimates of the vaccine efficacy against persistent infection with HPV 16 and 18 types (colours). Efficacy is shown over time with results from the included randomised controlled trials (indicated by shapes). B) The geometric mean antibody binding concentrations over time reported in each of the RCTs and against HPV 16 and 18. C) Relationship between antibody concentrations and vaccine efficacy when matching estimates from the same trials by time point and HPV types. The fitted relationship between efficacy and antibody binding concentrations is shown by the black solid line, with 95% CI (shaded region). Error bars: 95% credible intervals. Low opacity indicates less than four cases were reported in the control arm during the respective time interval.

### Using antibody binding and protection against other HPV types to identify a correlate of protection

The above analysis only included vaccine efficacy and antibody data for the HPV types 16 and 18, because these types where we had binding antibody data defined international units. However, five trials using Cervarix (bivalent vaccine), and one Gardasil-9 (nonavalent vaccine) trial reported vaccine efficacy against other HPV types (i.e. 31, 33, 45, 52, 58). The aggregated estimates of vaccine efficacy (by regimen) against these other HPV types are shown in Figure 2A. Gardasil-9 directly targets all these HPV types, and had correspondingly high efficacy against each type (Figure 2A). However, for the Cervarix vaccine, these HPV types are not targeted by the vaccine and protection to these types was lower (Figure 2A). We refer to HPV types not targeted by the vaccine as ‘off-types’ and protection against off-types is referred to as ‘cross-protection’. The range of observed cross-protection to these HPV types (31, 33, 45, 52, 58) highlights an opportunity to explore correlates of protection. However, to include these vaccine efficacy observations in a correlate of protection analysis, we needed to obtain binding antibody concentrations against these other HPV types. To do this, we utilised our previous search to identify all studies (not only RCTs) that contained immunogenicity data after Cervarix or Gardasil-9 vaccination (details in the supplementary methods) and identified one study of Cervarix vaccination^30^ and one study of Gardasil-9 vaccination^31^ that reported antibody concentrations against HPV types 31, 33, 45, 52 and 58. From these studies, we calculated the normalised binding concentration to each of the HPV types of interest from these studies (i.e. types 31, 33, 45, 52 and 58) relative to HPV16 (Figure 2B). We next scale the HPV16 concentrations reported in each RCT by the normalised antibody binding concentrations for these additional HPV types (Figure 2B), which gives estimates of the antibody concentrations to each of the HPV types after vaccination with Cervarix vaccine and Gardasil-9 (Table S7). In addition, five of the included RCTs reported immunogenicity against HPV 6 and 11 (though not in international units). Thus, for these types we followed the same procedure. That is, we normalised the reported HPV 6 and 11 geometric mean binding concentrations by the observed binding concentration to HPV16 (before conversion to international units) within the same study, to obtain the fold-difference between these types and HPV 16 (at each timepoint). We then multiplied these fol-differences by the corresponding trial specific HPV 16 binding concentration in IU/mL. We refer the antibody binding concentrations for these HPV types (6, 11, 31, 33, 45, 52 and 58) scaled to HPV 16 as being in ‘scaled international units’ per ml (sIU/mL). We match these antibody concentrations for each vaccine, regimen, HPV type and time since vaccination with the corresponding vaccine efficacy from each trial.

**Figure 2:**
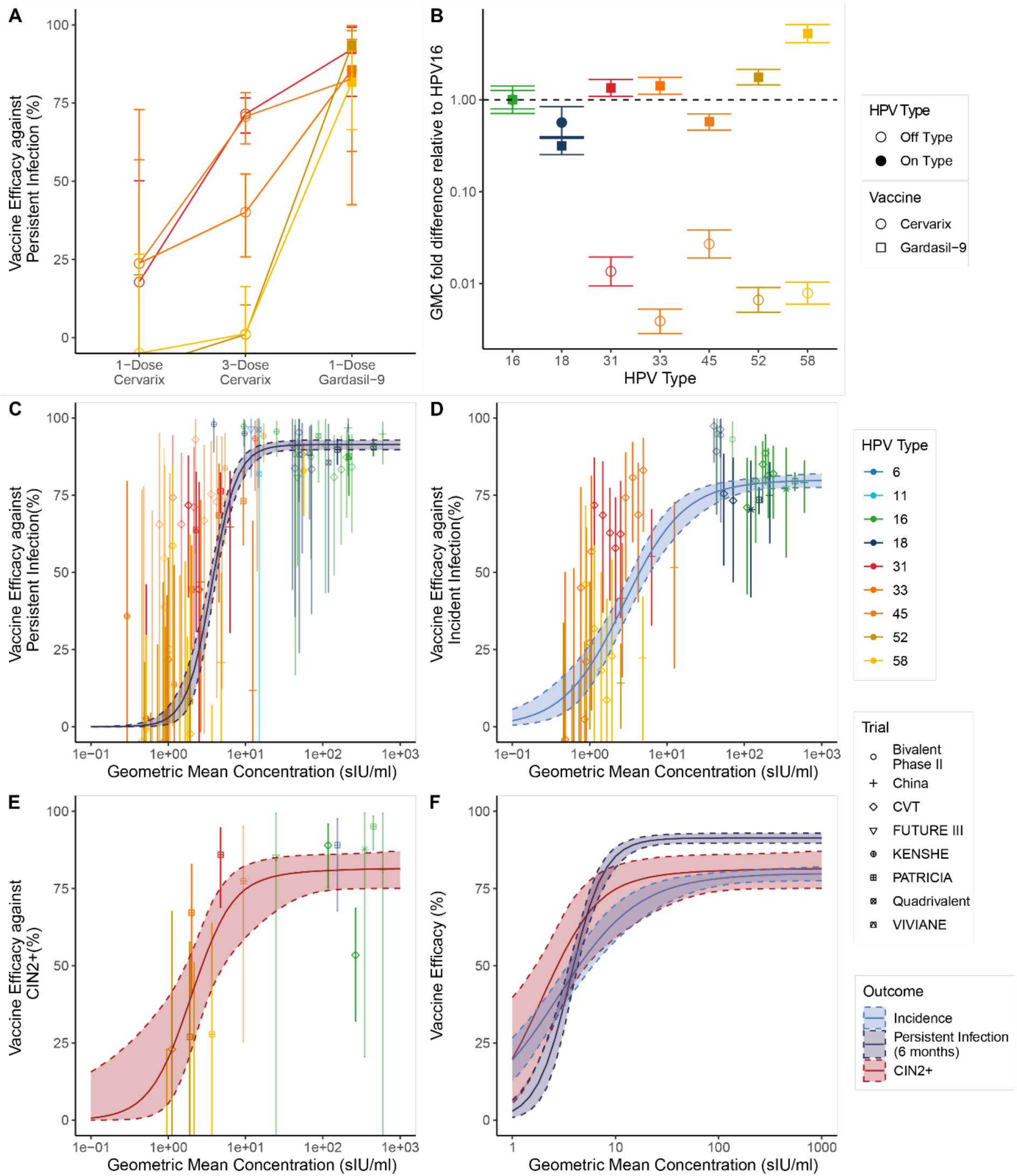
Vaccine efficacy and immunogenicity against all reported HPV types. A) Estimates of the vaccine efficacy against persistent infection with different HPV types (colour) (and 95% CI) provided by different vaccines from the included randomised controlled trials. B) Comparison of the geometric mean antibody concentrations for one and three doses of Cervarix and one dose Gardasil (shape) against the different HPV types. C-E) The relationship between HPV type-specific antibody binding concentrations and type-specific vaccine efficacy against C) persistent infection (*k* = 6.2 (CI:4.9-8.3)), D) incident infection (*k* = 2.5 (CI:1.8-3.5)), and E) CIN2+ (*k* = 3.7 (CI:1.6-6.5)). The solid lines indicate the fitted model associating antibody binding and vaccine efficacy for each of the three clinical outcomes and shaded regions indicate the 95% credible intervals. The clinical trials are indicated by different shapes, and observations with less than four cases in the control arm have reduced opacity. F) Comparison of the different fitted logistic curves for the different clinical outcomes. Error bars indicate 95% credible intervals. Scaled international units per ml (sIU/mL) indicates that we report antibody binding concentrations for HPV 16 and 18 in standard international units, and for other types we use the antibody binding concentration normalised to the HPV 16 concentration for each clinical trial (and so have been scaled to HPV 16 IU/mL).

Fitting all the available data (including, all HPV types and vaccine regimens), we observe a relationship between antibody binding concentrations and vaccine efficacy (Figure 2C-E). We find that binding antibody concentrations against each HPV type measured after vaccination are strongly associated with vaccine efficacy against incident HPV, persistent HPV infections and CIN2+ with the corresponding HPV type (i.e.: *p* < 0.0005, for all comparisons, Figure 2C-E, Table 1). Interestingly, the fitted relationships between antibody binding and vaccine efficacy were very similar for each of the three HPV-related outcomes considered, with very similar EC_50_ and slope parameters (Figure 2F, Table 1). The main difference observed was the estimated maximum vaccine efficacy against each of the three outcomes (highest for persistent infection, 91% and lowest for incident infection, 80%, Table 1). Using a leave-one-out analysis, we find that results are robust to the exclusion of any one study (Table S8). Further, the results were consistent with non-per-protocol observations from the included studies that were not included in our main analysis (Figure S12). Together these analyses demonstrate that antibody binding concentration to different HPV types after vaccination are correlated with protection against infection, persistent infection and CIN2+ associated with those corresponding HPV types.

**Table 1:** Estimates of the maximum efficacy, slope of logistic function and antibody binding concentration associated with 50% of the maximum efficacy obtained from the fitted model relating binding antibodies to vaccine efficacy in seronegative subjects. Note when k>0 this indicates a positive association between antibody binding and vaccine efficacy. Scaled international units per ml (sIU/mL) indicates that we report antibody binding concentrations for HPV 16 and 18 in standard international units, and for other types we use the antibody binding concentration normalised to the HPV 16 concentration for each clinical trial (and so have been scaled to HPV 16 IU/mL).

| Clinical Outcome reported | Estimated Maximum Efficacy | Slope of logistic function, k | Estimated concentration that gives 50% of the maximum level of protection (EC<sub>50</sub>, sIU/ml) |
| --- | --- | --- | --- |
| Incident HPV infection | 79.9% (78-83%) | 2.5 (1.8-3.5) | 2.8 (2.1-3.7) |
| Persistent HPV Infection (6 months) | 91.4% (89.7-93.0%) | 6.2 (4.9-8.3) | 3.6 (3.3-4.0) |
| CIN2+ | 81.4% (75-87%) | 3.7 (1.6-6.5) | 2.0 (1.2-3.5) |

### Comparison with protection against incident infection observed after natural infection

Several of the included RCTs followed individuals who were seropositive at baseline and not vaccinated, and in exploratory analyses, these showed associations of seropositivity with reduced risk of incident HPV infection. In a systematic review by Yokoji et al.,^32^ 25 studies were identified that compared risk of incident HPV infection (persistent infection and CIN2+ outcomes were not included) in seropositive and seronegative subjects. The GMC of HPV16-specific antibodies for seropositive (unvaccinated, presumed naturally infected) individuals at baseline of the CVT study was 2.8 sIU/ml^7^, which in our model equates to 40% protection (CI: 34-45), which is equivalent to a relative risk of 0.60 (CI: 0.55-0.66) (i.e. one minus vaccine efficacy gives the relative risk, Figure 3A-B). Aggregating the studies identified by Yokoji et al. the authors estimated that the relative risk of 0.70 (CI: 0.61-0.80) for HPV infection between seropositive and seronegative individuals (Figure 3C), which is similar to our prediction 0.60 (CI: 0.55-0.66).

**Figure 3:**
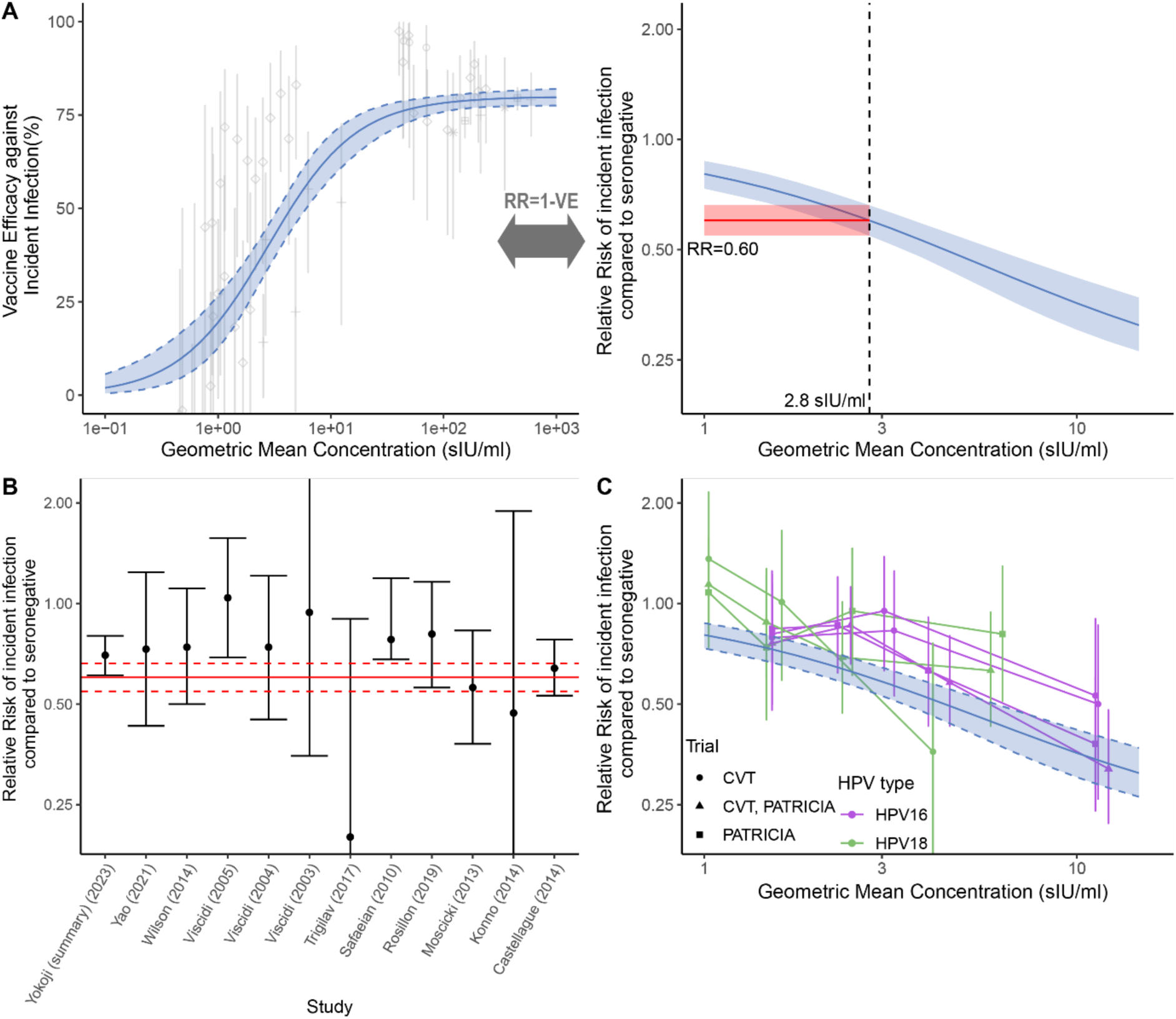
Comparing protection from vaccination and natural infection. A) Converting the relationship between antibody binding concentrations and vaccine efficacy against incident infection (from Figure 2D) to a relationship between antibody binding concentrations and the relative risk between seropositive and seronegative individuals (this curve is also used in panels B and C). Seropositive individuals had an antibody binding concentration of 2.8 sIU/ml to HPV16 on average in the CVT trials^7^, and at this concentration we predict a relative risk of 0.60 of incident HPV infection in seropositive individuals compared to seronegative individuals (red line). B) Estimates of relative risk of incident HPV infection in seropositive individuals compared to seronegative individuals (95% CI indicated by bars) from natural history studies identified in an existing systematic review (from Yokoji et al^32^), including an overall estimate (bold). The solid red line indicates our model-estimated relative risk of seropositive individuals (using the protection after vaccination), compared to seronegative individuals (with 95% CI indicated by the red shaded region). C) The relationship between antibody concentration and risk of incident HPV infection in seropositive individuals within the control arms of the PATRICIA and CVT vaccine trials is shown in green and purple lines (identified by an existing systematic review from Yokoji et al.^32^). This is compared with relationship between antibody concentration and protection observed after vaccination predicted by our study (blue line and shaded 95% CI). Scaled international units per ml (sIU/mL) is equivalent to IU/ml for HPV types 16 and 18.

In addition, three studies from Yokoji et al^32^ split the seropositive cohort by their binding antibody concentration and reported infection rates for these subgroups. These studies were sub-analyses of control arms of the CVT and PATRICIA randomised trials (including one study that combined the trials) and only reported protection against incident infection with HPV16 and HPV18. We extracted this data to compare the relative risk observed for different levels of naturally acquired antibodies to the relative risk predicted by our correlate of protection using the respective antibody concentrations of these groups (Figure 3C). This comparison suggests that the relationship between antibody levels and protection from incident infection is very similar between naturally acquired and vaccine-induced antibodies.

### HPV-type specific correlates of protection

The analyses above aggregate data across different HPV types and demonstrate an association between type-specific antibody binding concentrations and vaccine efficacy. However, the extent to which antibody binding between HPV types is comparable is not clear. Here we perform an additional analysis to determine whether the observed correlation is still evident within each HPV type. We observe that antibody binding concentrations are significantly associated with vaccine efficacy against persistent infection for HPV types 31, 52 and 58 (Figure S13). For HPV types 33 and 45, we see a trend for an association but do not reach significance. For HPV16 and 18, since we only observe high vaccine efficacy, we cannot see evidence of an association, and there is insufficient data for HPV 6 and 11 (Figure S13). For incident HPV and CIN2+ outcomes there was insufficient data to assess correlations by HPV type (Figure S14, S15).

Having seen evidence of a correlation between binding and vaccine efficacy for the stratified HPV types (for the outcome of persistent infection), we next consider whether the correlate of protection for each HPV type differs. That is, although antibody concentrations are correlated with vaccine efficacy, it is possible that the binding concentration associated with 50% protection (EC50) for each type may vary by type (e.g. in pneumococcal vaccination there is evidence for different protective thresholds for each strain)^33^. Therefore, we refitted the entire data set with a model allowing the EC50s to differ between the different HPV types but keeping the slope and maximum efficacy equivalent across the HPV types. We find that, there is a correlation between binding concentration and efficacy, however, we estimate significantly different EC50s between HPV types (Figure 4, Table S9). In particular, the 50% protective concentration for HPV16 and 18 were estimated to be low (HPV16, 0.54 IU/ml (0.05,1.9) and HPV18, 0.28 IU/ml (0.02,1.06)), while the 50% protective concentration was estimated to be significantly higher for HPV 58 (16.7 sIU/ml CI: (9.8,29.8)), with other types in between. For incident HPV infection and CIN2+ outcomes, we had insufficient data to estimate type specific EC50s (Figure S14, S15). Together, these data suggest that although antibody binding and vaccine efficacy against persistent infection are consistently correlated with vaccine efficacy across multiple HPV types, the concentration required for a certain level of protection may differ by HPV type.

**Figure 4:**
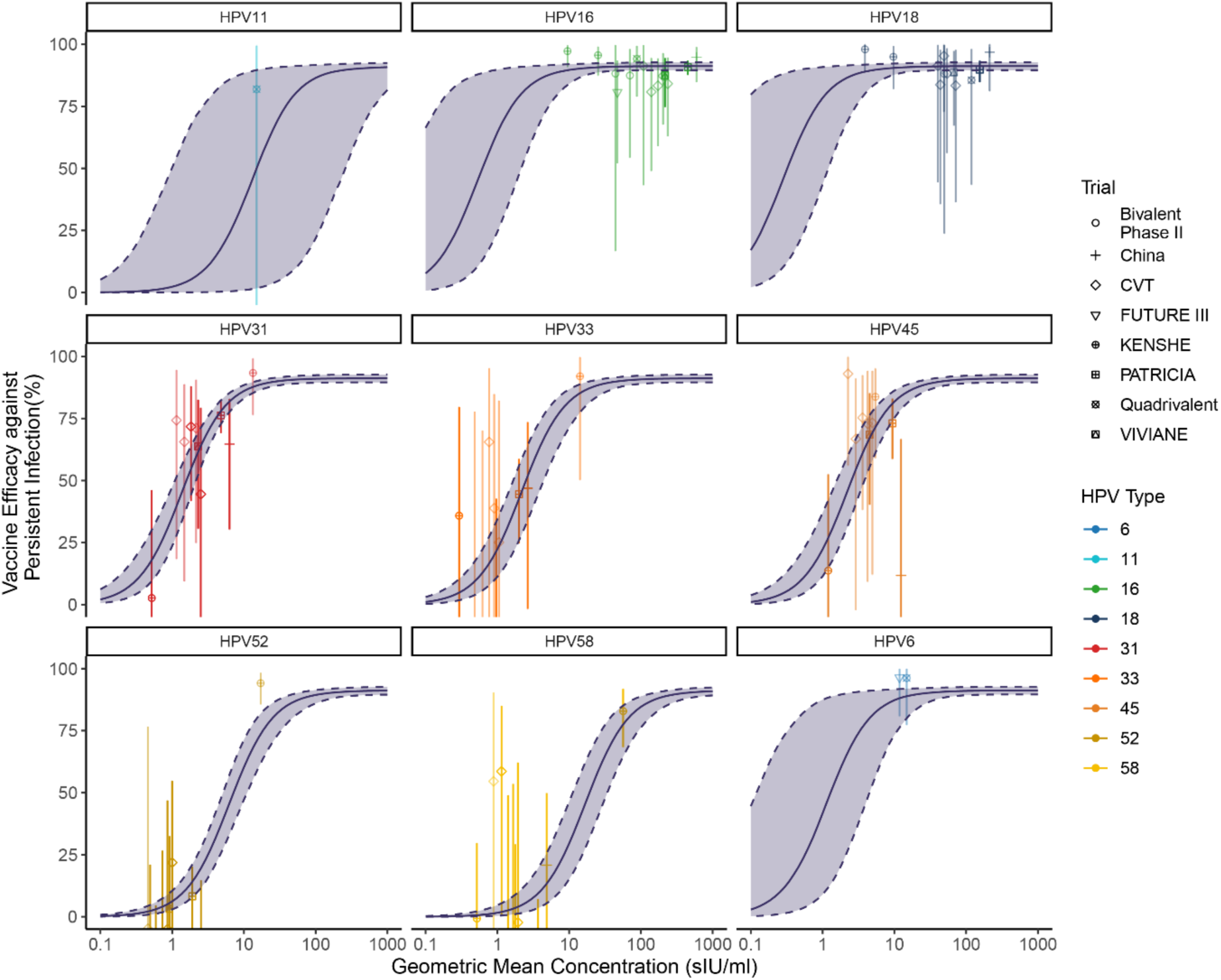
Type specific HPV correlates of protection. The relationship between binding antibodies and vaccine efficacy against persistent infection for each HPV type (colour) across all the trials (shape) allowing the EC50 to differ between each type but keeping the same maximum efficacy (*M* = 91.8, 95% CI: 90.2-93.1) and slope (*k* = 3.4, 95%CI: 2.7-4.1). Scaled international units per ml (sIU/mL) indicates that we report antibody binding concentrations for HPV 16 and 18 in standard international units, and for other types we use the antibody binding concentration normalised to the HPV 16 concentration for each clinical trial (and so have been scaled to HPV 16 IU/mL).

## Discussion

HPV-specific antibodies are thought to play a major role in vaccine-mediated protection^34^. However, to date, there has been no agreed correlate of protection for HPV that might be used for immunobridging to facilitate future vaccine assessment^14^. Several studies have reported an association between binding antibody concentrations and protection from incident infection in natural history studies^32,35^. In addition, studies have examined seropositivity and neutralising antibody titres to different HPV types and found associations with protection^36,37^. We have aggregated data across all RCTs that measured antibody binding and vaccine efficacy against HPV to understand the relationship between immunogenicity and protection. We find an association between HPV type-specific antibody binding after vaccination and clinical outcomes (across the reported outcomes of incident infection, 6-month persistent infection and CIN2+) (Figure 2). The evidence for this association arises primarily because of HPV types 31, 33, 45, 52 and 58. This is because there were relatively low observed efficacies to these HPV types particularly after one or three dose Cervarix vaccination. Even when each of these types are considered individually, we see evidence of positive associations between antibody binding and vaccine efficacy (only significant for 31, 52 and 58). Relatedly, it is important to notice that in similar stratified analyses for HPV 16 and 18, we do not see evidence of a correlation between antibody binding – because there were no observations of low vaccine efficacy against these HPV types. Nonetheless, in natural infection cohorts, we observe lower antibody levels and correspondingly lower protection against HPV 16, consistent with our analysis (Figure 3). Therefore, this work suggests that regardless of whether antibodies arose from HPV types included in the vaccine, from cross reactivity to non-vaccine HPV types, or after natural infection, antibody binding is, in general, associated with protection against HPV related outcomes.

We also find that the EC50s may differ between HPV types (this was only observed for protection against persistent infection – where sufficient data was available). This may be because of type-specific differences in infection (e.g. replicative fitness, pathogenesis etc). However, it may also be that differences in the EC50 arise due to differences in antibody binding assays across different types making them less comparable. In the case of the latter, the differences between EC50s for each type might be thought of as conversion factors between binding units for each type. Regardless, we observe consistently that antibody binding concentrations are associated with vaccine effectiveness, both within HPV types (e.g. stratification, Figure 4) and across HPV types (Figure 2). These observations support the use of antibody binding concentration as an endpoint in non-interiority trials to assess new vaccines.

A priority for global decision makers in recent years has been the decision of whether to switch to recommending a single dose HPV vaccination schedule, rather than two-dose schedules (of bivalent vaccines). Single-dose regimens have been adopted in high income and LMIC settings^38^, supported by clinical efficacy data^6^. However, some questions have persisted as to the durability of a single dose regimen. Since binding antibodies after one-dose of Cervarix vaccine are stable for at least 16 years, the correlation between antibody binding and vaccine efficacy observed here suggests that the observed efficacy will remain stable across that time period. Similarly, antibody concentrations after Gardasil-9 are also reported to be very stable over time across all antibody types,^31^ suggesting that the vaccine efficacy to each type will also be stable over time.

One strength of our analysis is the integration of data from a large number of studies. However, the included clinical studies also differed in a number of important respects, potentially impacting our comparisons between RCTs (Table S5). Although all of the RCTs we included in our analysis examined infection in women, they differed in age at vaccination (generally 15-25, but including one study of women 25-45 ^39^), duration of follow-up, and comparator group (some of the longer duration trials included secondary unvaccinated control groups^7^). Additionally, the trials are conducted in several different countries (Table S5). Further, the immunogenicity data obtained from each trial, is the geometric mean (usually of a subset of individuals in the trial) rather than individual antibody levels. Such differences introduce potential inconsistencies within our meta-analysis. However, our leave-one-out analysis suggests that these results were not strongly influenced by any one study (Table S8).

Another limitation of our study is the lack of a standardised HPV binding assay used across the studies. Our analysis combined results from assays that differed in format, antigen preparation and detection limits (which affected the definition of seropositivity, and therefore enrolment criteria, across studies). Two different assays were used across the different trials. Encouragingly, data extracted from Safaeian et al.^40^ indicates these two assays used in our analysis are well correlated (r=0.8, CI: 0.71-0.86) with a near 1:1 correspondence (Figure S16). A further study by Robbins et al.^41^ also found a high correlation between the two assays (*ρ* = 0.84 and *ρ* = 0.7, for HPV16 and HPV18 respectively). Further, to align assay differences as much as possible, in our primary analysis we used antibody concentrations from these assays reported in international units, or we converted the reported units to international units (supplementary methods S6). These conversion factors were sourced from the literature^21,22,40^(Table S4). The differences in the assays used should be taken into consideration when interpreting these results. A sensitivity analysis was conducted enabling different *EC*_50_ parameters for each assay and found that the correlation remained even after adjusting for differences in assays, and the *EC*_50_ parameters did not significantly differ between the assays (Table S10).

Finally, none of the identified RCTs report immunogenicity data for HPV types 31, 33, 45, 52 or 58. Thus, to include the vaccine efficacy data for these types in our analysis we incorporated data on antibodies against these HPV types from two separate studies (one for Cervarix and one for Gardasil-9), which were unrelated to the included RCTs. Thus, these two studies had different cohorts of vaccinated individuals and used different binding assay to the included RCTs^30,31^. We used this data to estimate the average fold difference in antibody binding concentration between these HPV types and HPV16. These fold differences were applied to the antibody data for each RCT to gain a specific estimate of antibody binding concentrations to each type for each RCT. An important limitation of this approach is that our estimates of the antibody binding concentration to HPV types 31, 33, 45, 52 and 58 are dependent on the two studies we identified that contained binding antibody data to these types after vaccination. It is possible that the fold differences between each HPV type and HPV 16 observed for Cervarix and Gardasil-9 would vary across studies (e.g. due to cohort and assay differences) if more data were available. Further examination of immunogenicity to these important oncogenic HPV types would strengthen analysis of correlates of protection. It is against these HPV types where the strongest evidence of a correlate of protection is observed (since it is against these HPV types where vaccine efficacies are observed that are not exclusively very high).

Despite these limitations, encouragingly, another study has used a neutralisation assay to show that neutralising antibodies to each HPV type is associated with vaccine efficacy to each type in the CVT and FUTURE trials^36^, consistent with our observations. Reanalysing this from Mariz et al.^36^ shows a similar relation to that which was identified here for binding antibody concentration (Table S11). None-the-less, a greater emphasis on antibody binding to HPV types other than 16 and 18 within clinical trials would strengthen assessment of this biomarker as a correlate of protection.

The ideal correlate of protection assay would be robust, standardised, high throughput and highly predictive of efficacy^42,43^. Binding antibody assays may not capture differences in antibody affinity and subsequently may not reflect true differences in functional inhibition of each HPV type. Neutralisation assays may capture more of the functionally relevant differences in antibodies and may produce a more generalisable correlate across HPV types. However, neutralising antibody assays are usually less high-throughput and less easily standardised. Functions other than neutralisation, such as Fc related functions, and cellular immune function (including T-cell mediated killing) may also play important roles in controlling HPV infection (particularly against long-term outcomes)^44,45^. However, there was very limited data to explore these biomarkers, these assays are typically difficult to standardise, and they often lack a high throughput format. These factors limit their utility as correlate of protection for decision making purposes.

Although our analysis comes with a number of caveats, the observed association between antibody binding and protection across vaccines, number of doses, HPV types, exposure histories, and time suggests that antibody binding may provide a useful surrogate marker for HPV vaccine effectiveness. Given the high efficacy of HPV vaccines, establishing non-inferiority of new vaccines or vaccine regimens based on clinical outcomes is likely to require very large studies. Our analysis suggests that establishing non-inferiority of antibody binding concentrations and/or neutralising antibody titres against existing vaccines may provide an alternative means of establishing effectiveness.

Here we have shown that antibody binding concentrations after HPV vaccination are correlated with protection against HPV infection and related clinical outcomes. The evidence of antibody responses after vaccination being a consistent and robust correlate is strengthened by comparing our predictions to the existing body of literature on correlates of protection in HPV. This work presents a strong basis for the use of antibody binding data to inform vaccine assessment and policy decisions into the future.

## Data Availability

Code and extracted data will be made publicly available on GitHub after publication.

## Contributors

MTB - Concept development, data curation, formal analysis, investigation, methodology, visualisation, writing manuscript. MRP - Concept development, data curation, investigation, methodology, writing manuscript. SRK - data curation, methodology, reviewing & editing EBR - data curation, methodology, reviewing & editing. EE - data curation, methodology, reviewing & editing. MAS - Concept development, supervision, review and editing DB - Concept development, review and editing. KC - Concept development, funding acquisition, supervision, review & editing. MPD - Concept development, funding acquisition, supervision, writing manuscript. DSK - Concept development, investigation, supervision, writing manuscript. All authors have read and approved the final version of the manuscript.

## Acknowledgements

This work is supported by National Health and Medical Research Council of Australia Investigator Grants (GNT1173027 to MPD, GNT2033318 to DSK) and Australia Leadership Investigator grants (APP1194679 to KC). DSK is supported by a University of New South Wales Scientia Fellowship.

## Data Sharing Statement

Code and extracted data will be made available publicly available after publication.

## References

1. Wei, F., Georges, D., Man, I., Baussano, I. & Clifford, G.M. Causal attribution of human papillomavirus genotypes to invasive cervical cancer worldwide: a systematic analysis of the global literature. Lancet 404, 435–444 (2024).

2. Schiffman, M., Clifford, G. & Buonaguro, F.M. Classification of weakly carcinogenic human papillomavirus types: addressing the limits of epidemiology at the borderline. Infect Agent Cancer 4, 8 (2009).

3. Bouvard, V., et al. A review of human carcinogens-Part B: biological agents. The Lancet Oncology 10, 321–322 (2009).

4. IARC. Cervical Cancer Screening, (IARC Handbook on Cancer Prevention, 2022).

5. Clifford, G.M., et al. Human Papillomavirus Genotype Distribution in Low-Grade Cervical Lesions: Comparison by Geographic Region and with Cervical Cancer. Cancer Epidemiology, Biomarkers & Prevention 14, 1157–1164 (2005).

6. Barnabas Ruanne, V., et al. Efficacy of Single-Dose Human Papillomavirus Vaccination among Young African Women. NEJM Evidence 1, EVIDoa2100056 (2022).

7. Kreimer, A.R., et al. Evaluation of Durability of a Single Dose of the Bivalent HPV Vaccine: The CVT Trial. J Natl Cancer Inst 112, 1038–1046 (2020).

8. Villa, L.L., et al. Prophylactic quadrivalent human papillomavirus (types 6, 11, 16, and 18) L1 virus-like particle vaccine in young women: a randomised double-blind placebo-controlled multicentre phase II efficacy trial. Lancet Oncol 6, 271–278 (2005).

9. Lei, J., et al. HPV Vaccination and the Risk of Invasive Cervical Cancer. New England Journal of Medicine 383, 1340–1348 (2020).

10. Jomah, A. & Albokhary, A. The impact of the human papillomavirus vaccine on women’s health: a systematic review. Eur Rev Med Pharmacol Sci 28, 3871–3879 (2024).

11. Palmer, T.J., et al. Invasive cervical cancer incidence following bivalent human papillomavirus vaccination: a population-based observational study of age at immunization, dose, and deprivation. JNCI: Journal of the National Cancer Institute 116, 857–865 (2024).

12. Falcaro, M., Soldan, K., Ndlela, B. & Sasieni, P. Effect of the HPV vaccination programme on incidence of cervical cancer and grade 3 cervical intraepithelial neoplasia by socioeconomic deprivation in England: population based observational study. BMJ 385, e077341 (2024).

13. WHO. Human Papillomavirus vaccines:WHO position paper (2022 update). Vol. 50 645–672 (2022).

14. Lehtinen, M., et al. Scientific approaches to defining HPV vaccine-induced protective immunity. International Journal of Cancer 156, 1848–1857 (2025).

15. Findlow, J., Lucidarme, J., Taha, M.-K., Burman, C. & Balmer, P. Correlates of protection for meningococcal surface protein vaccines: lessons from the past. Expert Review of Vaccines 21, 739–751 (2022).

16. Khoury, D.S., et al. Neutralizing antibody levels are highly predictive of immune protection from symptomatic SARS-CoV-2 infection. Nat Med 27, 1205–1211 (2021).

17. Berry, M.T., et al. Predicting vaccine effectiveness for mpox. Nat Commun 15, 3856 (2024).

18. Arbyn, M., Xu, L., Simoens, C. & Martin-Hirsch, P.P.L. Prophylactic vaccination against human papillomaviruses to prevent cervical cancer and its precursors. Cochrane Database of Systematic Reviews (2018).

19. Bergman, H., et al. Comparison of different human papillomavirus (HPV) vaccine types and dose schedules for prevention of HPV-related disease in females and males. Cochrane Database of Systematic Reviews (2019).

20. Henschke, N., et al. Efficacy, effectiveness and immunogenicity of one dose of HPV vaccine compared with no vaccination, two doses, or three doses. (Cochrane Response, 2022).

21. Tsang, S.H., et al. Evaluation of serological assays to monitor antibody responses to single-dose HPV vaccines. Vaccine 38, 5997–6006 (2020).

22. Nolan, K.M., et al. Development and Validation of Two Optimized Multiplexed Serologic Assays for the 9-Valent Human Papillomavirus Vaccine Types. mSphere 8, e00962–00921 (2023).

23. Kemp, T.J., et al. WHO International Standards for antibodies to HPV6 HPV11 HPV31 HPV33 HPV45 HPV52 and HPV58. npj Vaccines 9, 165 (2024).

24. Elias, K.M., et al. Viral clearance as a surrogate of clinical efficacy for COVID-19 therapies in outpatients: a systematic review and meta-analysis. The Lancet Microbe 5, e459–e467 (2024).

25. Stan Development Team. RStan: The R interface to Stan. (2023).

26. Stan Development Team. Stan Modeling Language Users Guide and Reference Manual. (2023).

27. R Foundation for Statistical Computing. R: A Language and Environment for Statistical Computing. (2024).

28. Koutsky, L.A., et al. A Controlled Trial of a Human Papillomavirus Type 16 Vaccine. New England Journal of Medicine 347, 1645–1651 (2002).

29. Zhao, F.H., et al. Efficacy, safety, and immunogenicity of an Escherichia coli-produced Human Papillomavirus (16 and 18) L1 virus-like-particle vaccine: end-of-study analysis of a phase 3, double-blind, randomised, controlled trial. Lancet Infect Dis 22, 1756–1768 (2022).

30. Pasmans, H., et al. Long-term HPV-specific immune response after one versus two and three doses of bivalent HPV vaccination in Dutch girls. Vaccine 37, 7280–7288 (2019).

31. Kemp, T.J., et al. Immunogenicity of one and two doses of Gardasil®9 in Tanzanian girls in the DoRIS Trial. npj Vaccines 11, 22 (2025).

32. Yokoji, K., et al. Association of naturally acquired type-specific HPV antibodies and subsequent HPV re-detection: systematic review and meta-analysis. Infect Agent Cancer 18, 70 (2023).

33. Andrews, N.J., et al. Serotype-specific effectiveness and correlates of protection for the 13-valent pneumococcal conjugate vaccine: a postlicensure indirect cohort study. The Lancet Infectious Diseases 14, 839–846 (2014).

34. Schiller, J. & Lowy, D. Explanations for the high potency of HPV prophylactic vaccines. Vaccine 36, 4768–4773 (2018).

35. Safaeian, M., et al. Risk of HPV-16/18 Infections and Associated Cervical Abnormalities in Women Seropositive for Naturally Acquired Antibodies: Pooled Analysis Based on Control Arms of Two Large Clinical Trials. J Infect Dis 218, 84–94 (2018).

36. Mariz, F.C., et al. Sustainability of neutralising antibodies induced by bivalent or quadrivalent HPV vaccines and correlation with efficacy: a combined follow-up analysis of data from two randomised, double-blind, multicentre, phase 3 trials. The Lancet Infectious Diseases 21, 1458–1468 (2021).

37. Draper, E., et al. A Randomized, Observer-Blinded Immunogenicity Trial of Cervarix® and Gardasil® Human Papillomavirus Vaccines in 12-15 Year Old Girls. PLOS ONE 8, e61825 (2013).

38. Umutesi, G., et al. A framework policy analysis of single-dose HPV vaccination adoption in East Africa: a rapid review. BMC Health Services Research 26, 608 (2026).

39. Zhu, F.C., et al. Safety and immunogenicity of human papillomavirus-16/18 AS04-adjuvanted vaccine in healthy Chinese females aged 15 to 45 years: a phase I trial. Chin J Cancer 30, 559–564 (2011).

40. Safaeian, M., et al. Direct Comparison of HPV16 Serological Assays Used to Define HPV-Naïve Women in HPV Vaccine Trials. Cancer Epidemiology, Biomarkers & Prevention 21, 1547–1554 (2012).

41. Robbins, H.A., et al. Comparison of Antibody Responses to Human Papillomavirus Vaccination as Measured by Three Assays. Frontiers in Oncology Volume 3 - 2013(2014).

42. Plotkin, S.A. & Gilbert, P.B. Nomenclature for Immune Correlates of Protection After Vaccination. Clinical Infectious Diseases 54, 1615–1617 (2012).

43. Plotkin, S.A. Immunologic correlates of protection induced by vaccination. The Pediatric Infectious Disease Journal 20, 63–75 (2001).

44. De Martino, S., et al. Role of innate immunity in tumor microenvironment of HPV-associated anal cancer: The hypothetical beneficial role of γδ T cells. Critical Reviews in Oncology/Hematology 212, 104771 (2025).

45. Li, Y., Deng, J., Liu, Y. & Yu, S. HPV infection and the immune microenvironment in cervical cancer. Frontiers in Immunology Volume 16 **-** 2025(2025).

