## Supplementary Material for "HPV binding antibodies are associated with protection: a meta-analysis"

#### Supplementary Methods

##### S1. Systematic review of existing literature

The over-arching aim of this study was to identify an association between antibody binding concentrations induced by HPV vaccination and protection against HPV-infection or HPV-related disease outcomes. To address this question, we aimed to systematically obtain studies that assess the immunogenicity and efficacy of HPV vaccines. This was completed in several steps and is outlined in detail below:

*1. Scoping review*

Initially, a scoping review was conducted to identify existing RCTs and systematic reviews of RCTs that:

(a) reported measures of HPV vaccine efficacy,

(b) reported measures of immunogenicity,

(c) included males or females age >9 years, and/or

(d) examined participants that received at least one-dose of an HPV vaccine.

Briefly, from this scoping review we identified the early FUTURE I, II and III group of RCTs which evaluated the efficacy of the 4vHPV vaccine (Table S12). The KENSHE study was a very large placebo-controlled, blinded trial evaluating persistent cervical HPV infection as the primary outcome measure, allowing direct measurement of vaccine efficacy[1]. The CVT and IARC India HPV trial were both RCTs that reported a large sample size[2,3]. KENSHE, DoRIS, CVT, and IARC India trial reported blinded laboratory measures of vaccine efficacy and immunogenicity. The PATH – Single dose HPV vaccine consortium also reported measures of efficacy and immunogenicity data from RCTs and observational studies comparing 1-dose, and 2 or 3-dose vaccine schedules[4].

We also identified three Cochrane systematic reviews and meta-analyses that reported efficacy and immunogenicity data for the 2vHPV, 4vHPV, and 9vHPV vaccines[5-7]. The earliest of the Cochrane reviews (2018)[5] evaluated prophylactic HPV vaccination to prevent precancerous lesions, and cervical cancer. A more recent Cochrane review (2019)[6] compared different HPV vaccine types and dose schedules for the prevention of HPV related disease in both males and females and evaluated the efficacy and immunogenicity of different vaccine dose schedules. The most recent Cochrane review (2022)[7] was an updated review which evaluated the efficacy, effectiveness, and immunogenicity with presumed previous infections, and 1, 2, or 3-dose schedules.

*2. Defining inclusion/exclusion criteria and search strategy*

Given the large number of available high quality RCTs of HPV vaccines reporting both antibody binding concentrations after vaccination as well as estimates of vaccine efficacy, we opted to focus our systematic review on RCTs only. We defined a set of study inclusion criteria outlined in Table S2.

The Cochrane reviews (Table S12) included studies that contained data on both the immunogenicity and efficacy of an HPV vaccines. Therefore, given the overlapping purposes of the Cochrane reviews and our own search we opted to utilise these Cochrane systematic reviews to identify studies that met our inclusion criteria. Our search strategy involved screening both included and excluded studies listed in the all the Cochrane reviews[5,7] for inclusion in our analysis and separately updating the 2022 Cochrane search to capture more recent studies.

*3. Updated literature review*

An update of the most recent Cochrane search published in 2022 was conducted. The 2022 Cochrane review included articles up to the 7 January 2022 inclusively. Therefore, we ran an updated search from this date up to 1st January 2026. Using the Central database, we searched MEDLINE (PubMed) and EMBASE (OVID) using the search terms provided in Table S1.

*4. Screening*

Studies identified by the Cochrane reviews (n=236, including 15 duplicate studies) and studies identified in our updated search of the literature (n=603, including 23 duplicates) underwent a title and abstract screening (excluded 646) then a full-text review against the inclusion criteria (excluded 92) following the criteria in Table S2. Papers were screened independently by two reviewers (from a team of four MTB, MRP, SRK and EE) with conflicts resolved by discussion amongst the four reviewers. This identified 87 papers that were included in our search relating to a total of 16 RCTs (Figure S1).

*5. Extraction*

Of the 87 papers identified for inclusion; it was observed that many papers reported data from the same clinical trials. Thus, to keep track of which data was from which clinical trial, during extraction each paper and RCT were assigned unique identifiers. One reviewer extracted data and a second reviewer, who was not the first reviewer, cross-checked the extracted data. The data extraction team was the same as the reviewer team (MTB, MRP, SRK and EE) All data was extracted into an extraction template that was separated in multiple levels. 1. Study, 2. Paper, and 3. Data (Table S3, Figure S17).

*6. Identifying most disaggregated data for inclusion in model-based meta-analysis*

Given that many publications were present for some RCTs, and to avoid using duplicated data in our meta-analysis, we chose to use only the most disaggregated (and non-overlapping) reports of immunogenicity and vaccine efficacy by time and HPV genotype that was available for each RCT. The papers included in our model-based meta-analysis are outlined in Table S5. For the FUTURE I & II studies, many of the efficacy data was pooled between the two trials, but the immunogenicity data was separate. This resulted in much of the data not being linked between the efficacy and immunogenicity.

In four trials[8-11], no disaggregation was made between outcomes related to HPV16 and HPV18, and instead a combined case count was reported. In these trials, to relate efficacy to immunogenicity, we assume that the reported case counts are indicative of HPV16 as this is the more common HPV type, and therefore we anticipate most cases to be related to HPV16 when the two HPV types are combined. Similarly, in one trial[12], no disaggregation was made between HPV6 and HPV11, and we use the reported case counts as representative of HPV6.

For the immunogenicity data, typically the extracted data was from the most recent publication as it contained the reported immunogenicity from the beginning of the trial. All extracted data used as part of the analysis is available in the GitHub repository, in the file 'Clinical_Trials_Data.xlsx' within the data folder.

*7. Rescreening for non-vaccine type immunogenicity*

To identify immunogenicity data for non-vaccine types we rescreened the excluded articles to identify potential studies that measured antibody binding after vaccination. From the excluded studies we identified three studies[13,14] that measured immunogenicity to non-vaccine HPV types. Of these three studies, one measured the effect of a Cervarix boost after primary vaccination with Gardasil[15]. However, since the time between vaccination and study enrolment was not recorded, this study was not able to be used.

#### S2. Modelling vaccine efficacy

To model vaccine efficacy (VE), we apply the same Bayesian methodology as previously reported[16]. We model the infection data using a binomial distribution where,

$$n_{(u,t)} \sim \mathrm{Bin}\left(r_{(u,t)}, N_{(u,t)}\right), \tag{1}$$

$$n_{(v,t)} \sim \mathrm{Bin}\left(r_{(u,t)}\left(1 - VE_{(v,t)}\right), N_{(v,t)}\right), \tag{2}$$

where, $n_{(u,t)}$ and $n_{(v,t)}$, are the number of events of a given outcome in unvaccinated and vaccinated groups, at time $t$, respectively. Also, $N_{(v,t)}$ is the number of participants in the vaccine arm, at time t, $N_{(u,t)}$ is the number of participants in the unvaccinated arm, at time t, $r_{(u,t)}$, is the infection probability in unvaccinated populations at time $t$, and $VE_{(v,t)}$ is the vaccine efficacy (and $1 - VE$ is the relative risk). This is applied to the most disaggregated data available within each study.

Using this model, we can predict an efficacy for a given vaccine and HPV type by assuming a constant vaccine efficacy across all studies and time for each vaccine and HPV type (figure 2A). To estimate the parameters, we chose the weakly-informative priors; a uniform distribution for the infection probability, $r \sim U(0,1)$, and for the vaccine efficacy, $VE_{(v,t)} \sim U(-1,1)$. We allow the VE to be negative to capture instances where the vaccine will not be effective.

**S3. Modelling antibodies as a correlate of protection.**

To relate the geometric mean antibody concentration (GMC) to protection we use a logistic model as previously described for other correlates[16-18]. This logistic model relates vaccine efficacy to geometric mean antibody concentration in the each of the vaccinated cohorts, and time points and against matched HPV types. We use the logistic function,

$$\mathrm{VE}_{(\mathrm{i,t})} = \frac{\mathrm{M}}{1 + exp\left(-k\left(log_{10}\left(GMC_{(i,t)}\right) - 1.5\right) + A\right)}, \quad (3)$$

where $VE_{(i,t)}$ is the vaccine efficacy at time-point $t$, and where $i$ denotes the index for the data identifying the trial, study arm and HPV type, with $GMC_{(i,t)}$ denoting the corresponding GMC. M denotes the maximum efficacy, $k$ represents the slope of the logistic function and approximates the log-odds ratio for a 10-fold change in concentration, and A is a parameter that captures the antibody concentration associated with 50% of the maximum vaccine efficacy, i.e. $EC_{50} = 10^{-\frac{A}{k}+1.5}$. This allows the vaccine efficacy to change over time and in different studies according to the reported GMC.

Using this function for the vaccine efficacy, we can transform this into relative risk between different antibody concentrations. When comparing between a naïve population we use that the relative risk, $RR = 1 - VE$, where VE is the vaccine efficacy.

All data used to fit equations (1) – (3) is included in the file 'Clinical_Trials_Data.xlsx' within the data folder of the linked GitHub repository. This includes all data from the studies listed in the summary table S5.

**S4. Selecting prior distributions**

To estimate the parameters, we use the prior distributions,

$$M \sim \mathrm{Uniform}(0,1), \quad (4)$$

$$k \sim \mathrm{Normal}(0,5), \quad (5)$$

$$A \sim \mathrm{Logistic}(0,1). \quad (6)$$

To obtain sensible estimates for vaccine efficacy we must use sensible weakly informative priors. Comparing the posterior and prior distributions for each parameter, we observe that the posterior is largely informed by the data (Figure S18).

To justify the choice of priors, we use our logistical model (equation (3)), with the case $k = 0$, where we do not have a correlate and the vaccine efficacy is given by,

$$VE = \frac{M}{1 + \exp(A)}. \tag{7}$$

This is equivalent to,

$$VE = M \operatorname{logit}(A). \tag{8}$$

It follows that, given the logistic prior for $A$, $\operatorname{logit}(A)$ will follow a uniform distribution between 0 and 1. Whereas, a diffuse prior on A (such as a commonly used normal distribution, $\text{Normal}(0,5)$) would place a higher prior probability on the VE being at the extreme values of 0 or 1 (Figure S19, setting $M = 1$). Likewise, we use the non-informative uniform distribution as the prior for the maximum value M. For the slope parameter, $k$, we select use a weakly-informative normal distribution as a standard approach.

***Sensitivity of the Prior Distributions***

To understand the impact of the choice of prior distributions we conduct a posterior sensitivity analysis where we select different prior distributions. For the parameters $A$, and $M$, we can see a potential limitation from equation 8. In this case, we observe that the vaccine efficacy is the product of two uniform distributions and subsequently skews towards a lower vaccine efficacy. To account for this, we conduct a sensitivity analysis using a $\text{Beta}(10,1)$ distribution as a prior for the maximum M. This results in a much more uniform prior on the VE in equation 8 (Figure S19). However, we found that the parameter estimates were insensitive to this choice of prior (Table S13). Similarly for the parameter A, we conducted a sensitivity using a normal distribution and increasing the prior variance, and likewise found the results were insensitive to the prior choice (Table S14).

For the slope parameter $k$, we use a diffuse normal distribution. However, there is a potential limitation of this choice of prior. If the distribution is very wide, it may artificially inflate the probability that the vaccine efficacy is close to 0 or the maximum ($M$) (Figure S19). Alternatively, for narrow distributions, then our prior likelihood becomes too concentrated around 0 and can become too informative of our model parameters. To test the effect of our prior choice, we conduct a sensitivity, varying the width of the prior distribution between $\text{Normal}(0,1)$ and $\text{Normal}(0,20)$. For normal distributions between $\text{Normal}(0,5)$ and $\text{Normal}(0,20)$ the estimates for $k$ were insensitive to the width of the prior distribution (Table S15). The narrower $\text{Normal}(0,1)$, is more conservative with respect to our null hypothesis, and has greater influence on the posterior distribution (compared to a Normal(0,5) distribution) decreasing the estimates for the slope parameters (central estimates and credible intervals). However, all choices for the prior still results in a significant slope of all outcomes (Table S15).

**S5. Scaled IU/mL for HPV types 31, 33, 45, 52 and 58**

To use binding antibody data, we used two additional studies (which were not otherwise connected to an included RCT) to obtain immunogenicity data against HPV types 31, 33, 45, 52 and 58[13,14]. Rather than use the raw concentrations from these two studies directly (since they used different assays and populations to the included RCTs) we opted to consider the fold differences to these types. To calculate the fold-differences we use the equation,

$$FD_{(r,h,t)} = \frac{GMC_{(r,h,t)}}{GMC_{(r,16,t)}}, \tag{7}$$

where $FD_{(r,h,t)}$, denotes the fold-difference in binding concentration for vaccine regimen (vaccine and dose combination), $r$, between HPV type $h$ and HPV 16, at time $t$ and $GMC_{(r,h,t)}$, denotes the geometric mean antibody concentration for vaccine regimen, $r$, against HPV type $h$, at time $t$. After computing the fold-differences at each time point, we calculate the geometric mean fold-difference, $GMFD_{r,h}$ against each HPV type, $h$, and vaccine regimen, $r$, as the geometric mean across all time points, i.e.

$$GMFD_{r,h} = \left(\prod_{t} FD_{(r,h,t)}\right)^{\frac{1}{j}} \tag{8}$$

where $j$ is the number of time points. Finally, to calculate the estimated GMC for study $s$, for regimen, $r$, against HPV type $h$, at time $t$, we use the equation,

$$\widetilde{GMC_{(s,r,h,t)}} = GMFD_{r,h} \times GMC_{(s,r,16,t)} \tag{9}$$

where, $GMC_{(s,r,16,t)}$, is the GMC reported against HPV16 in study $s$, for vaccine regimen, $r$, after $t$ months post vaccination. Since this GMC is reported in IU/ml, by applying these fold-differences we obtain GMC in a scaled international units denoted sIU/ml.

For one of the studies (Pasmans et al.[13]) the reported GMC includes only those who were deemed seropositive. However, a number of individuals were considered seronegative, where their concentration was below a specified limit of detection. To account for these individuals, we assigned these individuals a concentration of half the limit of detection. The GMC for the total population of $N$ individuals, using the reported GMC ($GMC_n$) for the $n$ seropositive individuals is then given by,

$$GMC = \left((GMC_n)^n \left(\frac{LOD}{2}\right)^{N-n}\right)^{\frac{1}{N}}, \tag{10}$$

where, LOD indicates the limit of detection.

**S6. Modelling neutralisation titres as a correlate of protection.**

As part of a discussion around functional correlates of protection we analysed data from Mariz et al.[19] To relate the geometric mean neutralisation titre (GMT) to vaccine efficacy observed in Mariz et al.[19] we again use a logistic model (similar to equation 3). In this study, we obtained only observed vaccine efficacy (OVE) estimates, rather than individual case data between study arms. In this scenario we have the logistic model,

$$VE_{\mathrm{i}} = \frac{\mathrm{M}}{1 + exp(-k(log_{10}(GMT_i) - 1.5) + A)}, \tag{13}$$

where, $VE_i$ denotes the expected vaccine efficacy, $i$ denotes the index for the data identifying the trial and HPV type, M denotes the maximum efficacy, $k$ represents the slope of the logistic function (and approximates the log-odds ratio for a 10-fold change in concentration), and A is a parameter that captures information on the GMT associated with 50% of the maximal protection (see above). We use a weighted beta-proportional likelihood function to model the observed VE, using the expected VE (MVE), and an additional parameter, $\kappa$, which relates to the precision in the observed value, that is,

$$VE_i \sim \text{Beta-Prop}(MVE_i, \kappa) * w_i \tag{14}$$

where,

$$w_i = \frac{n\left(CI_{i,max} - CI_{i,min}\right)^{-\frac{1}{2}}}{\sum_{j=1}^{n}\left(CI_{j,max} - CI_{j,min}\right)^{-\frac{1}{2}}}, \tag{15}$$

and where $CI_{i,max}$ and $CI_{i,min}$ refer to the maximum and minimum of the confidence interval of the vaccine efficacy for the $i$th observation. This model restricts the observed vaccine efficacies to lie in the interval (0,1). However, one observed VE was less than 0 (-8.9%) and this was set to be 0.0001. Similarly, one observed VE was 100%, which was adjusted so that the VE=0.9999. Adjustments were not made to the confidence intervals as only the interval length is needed to calculate the weights. To estimate the parameters, we use the prior distributions,

$$M \sim Uniform(0,1), \tag{16}$$

$$k \sim Normal(0,5), \tag{17}$$

$$A \sim \text{Logistic}(0,1). \tag{18}$$

$$\kappa \sim \text{INV-Gamma}(50,350) \tag{19}$$

### Supplementary Figures and Tables

| **Search strategy** |
|---|
| #1. MeSH descriptor Papillomavirus Infections explode all trees |
| #2. MeSH descriptor Papillomaviridae explode all trees |
| #3. (HPV*) |
| #4. (human papillomavirus*) |
| #5. (human papilloma virus*) |
| #6. (#1 OR #2 OR #3 OR #4 OR #5) |
| #7. MeSH descriptor Papillomavirus Vaccines explode all trees |
| #8. (gardasil) |
| #9. (cervarix) |
| #10. (vaccin*) |
| #11. (immuni*) |
| #12. (#7 OR #8 OR #9 OR #10 OR #11) |
| #13. (#6 AND #12) |

**Table S1. Description of search terms used to update the systematic review.**

**Study inclusion criteria**

*Type of study*

- Restricted to only phase II and phase III randomised controlled trials (RCTs).
- A registered RCT or a study linked to a registered RCT (including post-hoc analyses of one or more RCTs).

*Intervention*

Vaccination with at least one dose of a prophylactic HPV vaccine.

*Participant*

Studies on female or male participants aged ≥ 9 years, that excluded in their analysis known HPV positive individuals.

*Outcome*

Restricted to studies capable of providing data on the immunogenicity and efficacy of an HPV vaccine against any HPV associated outcome (e.g. Infection, CIN) and any HPV type.

*Control*

Placebo group containing no active product or only the adjuvant of the HPV vaccine, without L1 VLP, or another irrelevant vaccine.

**Table S2. Detailed description of inclusion criteria.**

**Study extraction categories**

*Study level*

- A unique study identifier
- Number of participants
- Age range

*Paper level*

- A unique paper identifier.
- Year published.
- Author,
- Title, and
- Associated trial.

*Data level*

- Efficacy
    - Trial number (clinical trial identifier),
    - Outcome (clinical outcome: Incidence, PI – persistent infection, CIN2+ - CIN2 or worse),
    - Vaccine (Name of vaccine used),
    - Doses (Number of HPV vaccine doses used),
    - Set (The category for the efficacy data (Per protocol Set (PPS), Intention to treat (ITT), or Total vaccinated cohort (TVC)),
    - HPV type (HPV type associated with outcome),
    - Start (years since vaccine administered)
    - End (years since vaccine administered),
    - Cases (n, number of cases of outcome),
    - Population (N, Total population),
    - Person-years (Number of person-years accounted for),
    - Serostatus (Negative if the set is all seronegative at baseline, positive if the set is positive at baseline, or left blank if it includes both. HPV DNA status should always be negative),
    - LTFU (Long term follow-up),
    - Table/figure (table or figure data was extracted from), and
    - Unique paper ID.

- Immunogenicity
    - Trial number (clinical trial identifier),
    - Assay type (Assay used: ELISA, or otherwise),
    - Vaccine (Name of vaccine used),
    - Doses (Number of HPV vaccine doses used),
    - HPV type (HPV type associated with outcome),
    - Set (The category for the efficacy data (Per protocol Set (PPS), Intention to treat (ITT), or Total vaccinated cohort (TVC)),
    - Time since first vaccine dose (months),
    - GMC* (Reported Geometric mean antibody concentration),
    - GMC* LCI (Lower limit of GMC),
    - GMC* UCI (Upper limit of GMC),
    - Population (N, Number of samples taken), and
    - Unique paper ID.

**Table S3. Detailed description of extraction parameters.** All information connected to the study was recorded at the study level. All details were recorded and linked to the unique study identifier at the paper level (Each study may be linked to more than one paper). All extracted data from the paper was recorded at the data level. *Immune markers that used endpoints other than GMC, such as mean fluorescence index (MFI) or geometric mean titres were extracted as GMCs with the assay type used to differentiate the markers.

| Assay | HPV16 conversion factor | HPV18 conversion factor |
|---|---|---|
| ELISA[20] | 5.29 EU=1 IU | 6.81 EU = 1 IU |
| cLIA[21] | 10 mMU=1 IU | 7.9 mMU = 1 IU |

**Table S4:** Conversion factors for the two different assays into international units[20,21].

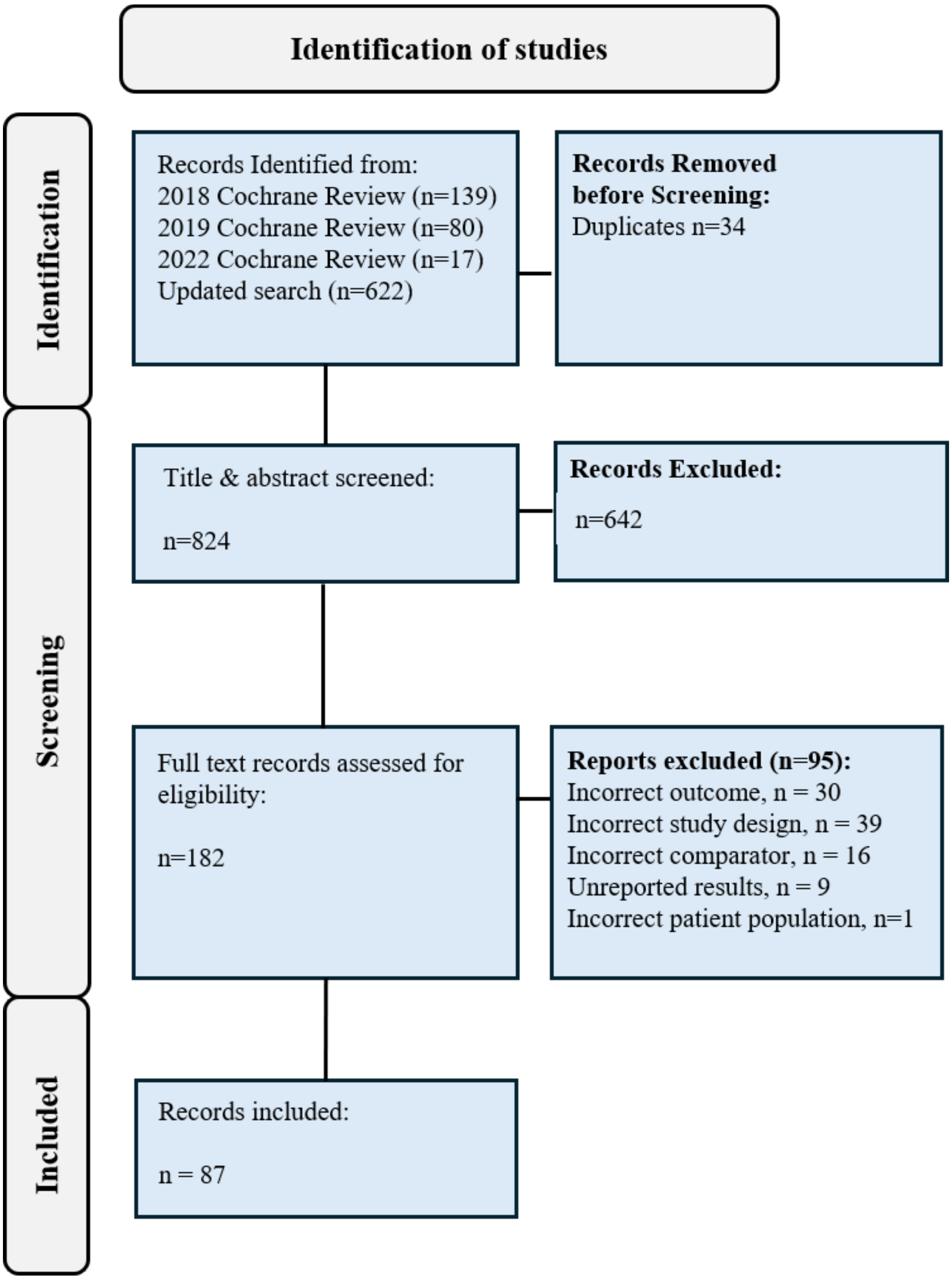

**Figure S1. PRISMA chart for the systematic review.** We identified records from three Cochrane reviews published in 2018, 2019 and 2022, and our own updated search from the Cochrane review end date 7 January 2022 to 1 January 2026. A summary of included studies is detailed in Tables S5 and S6.

| Trial (Trial Number) | Year started | Vaccine | Follow-up time (years) | Number of participants | Age Group | Clinical outcomes included in analysis | Immune Assay (timepoints) | Number of associated papers (used for extraction) |
|---|---|---|---|---|---|---|---|---|
| *CVT Study (NCT00128661)* | 2004 | Cervarix | 11 | 7466 | 18-25 | Incident infection<br>Persistent infection<br>Persistent infection (12m)<br>CIN2+ | ELISA (5) | 20† (3) |
| *Bivalent Phase II (NCT00689741)* | 2000 | Cervarix | 9 | 1113 | 15-25 | Incident infection<br>Persistent infection<br>Persistent infection (12m)<br>CIN2+ | ELISA (13) | 5 (4) |
| *China I (NCT01735006)* | 2012 | Cecolin | 5.5 | 8827 | 18-45 | Incident infection<br>Persistent infection<br>CIN2+ | ELISA (6) | 4 (1) |
| *China II (NCT00779766)* | 2008 | Cervarix | 6 | 3819 | 18-25 | Incident infection<br>Persistent infection<br>Persistent infection (12m)<br>CIN2+ | ELISA (3) | 6 (1) |
| *China III (NCT00834106)* | 2008 | Gardasil | 7.5 | 3006 | 20-45 | CIN2+ | cLIA (1) | 2 (1) |

| Trial (Trial Number) | Year started | Vaccine | Follow-up time (years) | Number of participants | Age Group | Clinical outcomes included in analysis | Immune Assay (timepoints) | Number of associated papers (used for extraction) |
|---|---|---|---|---|---|---|---|---|
| *Future I (NCT00092521)* | 2002 | Gardasil | 5 | 5455 | 16-24 | CIN2+ | cLIA (3) | 16#(2) |
| *Future II (NCT00092534)* | 2002 | Gardasil | 14 | 12167 | 15-26 | CIN2+ | cLIA (5) | 16# (2) |
| *Future III (NCT00090220)* | 2004 | Gardasil | 4 | 3819 | 24-45 | Persistent infection<br>CIN2+ | cLIA (5) | 4 (4) |
| *KENSHE (NCT03675256)* | 2018 | Cervarix<br>Gardasil-9 | 4.5 | 2275 | 15-20 | Persistent infection | ELISA (1) | 3 (2) |
| *Japan I* | 2006 | Cervarix | 4 | 1040 | 20-25 | Incident infection<br>Persistent infection<br>Persistent infection (12m)<br>CIN2+ | ELISA (6) | 3 (2) |

| Trial (Trial Number) | Year started | Vaccine | Follow-up time (years) | Number of participants | Age Group | Clinical outcomes included in analysis | Immune Assay (timepoints) | Number of associated papers (used for extraction) |
|---|---|---|---|---|---|---|---|---|
| *Japan II* | NA | Gardasil | 2.5 | 1021 | 18-26 | Persistent infection | cLIA (7) | 1 (1) |
| *Monovalent (NCT00365378)* | 1998 | Monovalent | 4 | 2392 | 16-23 | Incident infection<br>Persistent infection<br>CIN2+ | cLIA (9) | 3 (3) |
| *PATRICIA (NCT00122681)* | 2004 | Cervarix | 4 | 18644 | 15-25 | Incident infection<br>Persistent infection<br>Persistent infection (12m)<br>CIN2+ | ELISA (5) | 9† (3) |
| *Quadrivalent Phase II (NCT00365716)* | 2000 | Gardasil | 5 | 1158 | 16-23 | Persistent infection | cLIA (3) | 3 (1) |
| *VIVIANE (NCT00294047)* | 2006 | Gardasil | 7 | 5752 | >25 | Persistent infection<br>CIN2+ | cLIA (5) | 2 (2) |

| Trial (Trial Number) | Year started | Vaccine | Follow-up time (years) | Number of participants | Age Group | Clinical outcomes included in analysis | Immune Assay (timepoints) | Number of associated papers (used for extraction) |
|---|---|---|---|---|---|---|---|---|
| *Quadrivalent in Men (NCT00090285)** | 2004 | Gardasil | 3 | 4065 | 16-26 | No infection outcomes in analysis | cLIA (3) | 5 (3) |
| *UlacNet (NCT04255849)** | 2021 | Gardasil-9 | 4.5 | 700 | 20-50 | No infection outcomes in analysis | - | 1(1) |

**Table S5.** Summary of the Randomised Controlled Trials identified through our search, including which studies contained the most common clinical endpoints whilst additional endpoints were found in various trials, these are not listed in the table. * The quadrivalent in men and UlacNet studies were not included in the analyses as there were insufficient infection outcomes shared with other studies. † Three studies included data from both the CVT and PATRICIA trials. # 13 studies related to both the FUTURE I and FUTURE II studies. Persistent Infection= Two consecutive positive samples either four or 6 months apart, Persistent Infection 12m= At least two consecutive positive samples for at least 12 months without a negative sample. CIN2+= Cervical Intraepithelial Neoplasia grade 2 or higher, cLIA = competitive Luminex immunoassay, ELISA = Enzyme linked immunosorbent assay, LMS=Luminex multiplex serology, NA=not available.

| Author | Year | Title | Incident Infection | 6-month persistent infection | 12-month persistent infection | CIN2+ | Off-type efficacy | Immuno-genicity |
|---|---|---|---|---|---|---|---|---|
| <u>*CVT Study (NCT00128661)*</u> | | | | | | | | |
| Herrero[3] | 2011 | Prevention of persistent human papillomavirus infection by an HPV16/18 vaccine: a community-based randomized clinical trial in Guanacaste, Costa Rica | - | Yes | Yes | - | Yes | - |
| Kreimer[22] | 2011 | Efficacy of a bivalent HPV 16/18 vaccine against anal HPV 16/18 infection among young women: a nested analysis within the Costa Rica Vaccine Trial | Yes | - | - | - | - | - |
| Kreimer[23] | 2011 | Proof-of-principle evaluation of the efficacy of fewer than three doses of a bivalent HPV16/18 vaccine | - | Yes | Yes | - | - | - |
| Herrero[24] | 2013 | Reduced prevalence of oral human papillomavirus (HPV) 4 years after bivalent HPV vaccination in a randomized clinical trial in Costa Rica | Yes | - | - | - | - | - |

| Author | Year | Title | Incident Infection | 6-month persistent infection | 12-month persistent infection | CIN2+ | Off-type efficacy | Immuno-genicity |
|---|---|---|---|---|---|---|---|---|
| Safaeian[25] | 2013 | Durable antibody responses following one dose of the bivalent human papillomavirus L1 virus-like particle vaccine in the Costa Rica Vaccine Trial | - | - | - | - | - | Yes |
| Hildesheim[26] | 2014 | Efficacy of the HPV-16/18 vaccine: final according to protocol results from the blinded phase of the randomized Costa Rica HPV-16/18 vaccine trial | Yes | - | - | Yes | Yes | Yes |
| Lang Kuhs[27] | 2014 | Reduced Prevalence of Vulvar HPV16/18 Infection Among Women Who Received the HPV16/18 Bivalent Vaccine: A Nested Analysis Within the Costa Rica Vaccine Trial | Yes | - | - | - | - | - |
| Lang Kuhs[28] | 2014 | Effect of different human papillomavirus serological and DNA criteria on vaccine efficacy estimates | - | - | Yes | Yes | - | - |
| Beachler[9] | 2016 | Multisite HPV16/18 Vaccine Efficacy Against Cervical, Anal, and Oral HPV Infection | Yes | - | - | - | - | - |

| Author | Year | Title | Incident Infection | 6-month persistent infection | 12-month persistent infection | CIN2+ | Off-type efficacy | Immuno-genicity |
|---|---|---|---|---|---|---|---|---|
| Safaeian[29] | 2018 | Durability of Protection Afforded by Fewer Doses of the HPV16/18 Vaccine: The CVT Trial | Yes | - | - | - | - | Yes |
| Kreimer[30] | 2020 | Evaluation of Durability of a Single Dose of the Bivalent HPV Vaccine: The CVT Trial | - | - | - | - | - | Extracted |
| Tsang[31] | 2020 | Durability of Cross-Protection by Different Schedules of the Bivalent HPV Vaccine: The CVT Trial | Extracted | Extracted | - | - | Yes | - |
| Shing[32] | 2022 | Precancerous cervical lesions caused by non-vaccine-preventable HPV types after vaccination with the bivalent AS04-adjuvanted HPV vaccine: an analysis of the long-term follow-up study from the randomised Costa Rica HPV Vaccine Trial | - | - | - | Extracted | Extracted | - |
| Befano[33] | 2023 | Estimating human papillomavirus vaccine efficacy from a single-arm trial: proof-of-principle in the Costa Rica Vaccine Trial | - | - | - | - | - | - |

| Author | Year | Title | Incident Infection | 6-month persistent infection | 12-month persistent infection | CIN2+ | Off-type efficacy | Immuno-genicity |
|---|---|---|---|---|---|---|---|---|
| Wacholder[34] | 2010 | Risk of miscarriage with bivalent vaccine against human papillomavirus (HPV) types 16 and 18: pooled analysis of two randomised controlled trials | - | - | - | - | - | - |
| Kreimer[35] | 2015 | Efficacy of fewer than three doses of an HPV-16/18 AS04-adjuvanted vaccine: combined analysis of data from the Costa Rica Vaccine and PATRICIA Trials | Yes | Yes | Extracted | - | - | - |
| Kreimer[36] | 2018 | Evidence for single-dose protection by the bivalent HPV vaccine-Review of the Costa Rica HPV vaccine trial and future research studies | - | - | - | - | - | - |
| Harari[37] | 2016 | Cross-protection of the Bivalent Human Papillomavirus (HPV) Vaccine Against Variants of Genetically Related High-Risk HPV Infections | - | - | - | - | - | - |
| Haas[38] | 2025 | Design and initial findings of a natural history study of anal HPV and associated lesions among young adult women in Costa Rica | - | - | - | - | - | - |

| Author | Year | Title | Incident Infection | 6-month persistent infection | 12-month persistent infection | CIN2+ | Off-type efficacy | Immuno-genicity |
|---|---|---|---|---|---|---|---|---|
| Porras[39] | 2024 | HPV16/18 antibodies 16-years after single dose of bivalent HPV vaccination: costa Rica HPV vaccine trial | - | - | - | - | - | - |
| *Bivalent Phase II (NCT00689741)* | | | | | | | | |
| De Carvalho[40] | 2010 | Sustained efficacy and immunogenicity of the HPV-16/18 AS04-adjuvanted vaccine up to 7.3 years in young adult women | Yes | Yes | Yes | Yes | - | Extracted |
| Romanowski[41] | 2009 | Sustained efficacy and immunogenicity of the human papillomavirus (HPV)-16/18 AS04-adjuvanted vaccine: analysis of a randomised placebo-controlled trial up to 6·4 years | - | - | - | - | | Yes |
| Harper[42] | 2004 | Efficacy of a bivalent L1 virus-like particle vaccine in prevention of infection with human papillomavirus types 16 and 18 in young women: a randomised controlled trial | Extracted | Extracted | - | - | - | Yes |

| Author | Year | Title | Incident Infection | 6-month persistent infection | 12-month persistent infection | CIN2+ | Off-type efficacy | Immuno-genicity |
|---|---|---|---|---|---|---|---|---|
| Harper[43] | 2006 | Sustained efficacy up to 4.5 years of a bivalent L1 virus-like particle vaccine against human papillomavirus types 16 and 18: follow-up from a randomised control trial | Extracted | Extracted | Extracted | Extracted | - | |
| Naud[8] | 2014 | Sustained efficacy, immunogenicity, and safety of the HPV-16/18 AS04-adjuvanted vaccine: final analysis of a long-term follow-up study up to 9.4 years post-vaccination | Extracted | Extracted | Extracted | Extracted | - | - |
| Roteli-Martins[44] | 2012 | Sustained immunogenicity and efficacy of the HPV-16/18 AS04-adjuvanted vaccine: up to 8.4 years | - | - | - | - | - | Yes |
| *China I (NCT00779766)* | | | | | | | | |
| Zhao[45] | 2022 | Efficacy, safety, and immunogenicity of an Escherichia coli-produced Human Papillomavirus (16 and 18) L1 virus-like-particle vaccine: end-of-study analysis of a phase 3, double-blind, randomised, controlled trial | Extracted | Extracted | Extracted | Extracted | Extracted | Extracted |

| Author | Year | Title | Incident Infection | 6-month persistent infection | 12-month persistent infection | CIN2+ | Off-type efficacy | Immuno-genicity |
|---|---|---|---|---|---|---|---|---|
| Zhao[11] | 2025 | Long-term efficacy and immunopersistence of an Escherichia coli-produced HPV-16/18 bivalent vaccine: an observational extension study following a randomised, double-blind Phase III clinical trial cohort | - | Extracted | - | - | - | Extracted |
| Jia[46] | 2025 | Long-term protective efficacy of the Escherichia coli-produced HPV-16/18 bivalent human papillomavirus vaccine in women vaccinated at 18-45 years: a 9-year follow-up study | - | - | - | - | - | - |
| Su[47] | 2024 | Pattern of multiple human papillomavirus infection and type competition: an analysis in healthy Chinese women aged 18-45 years | - | - | - | - | - | - |
| *China II (NCT01735006)* | | | | | | | | |
| Zhu[48] | 2014 | Efficacy, immunogenicity and safety of the HPV-16/18 AS04-adjuvanted vaccine in healthy Chinese women aged 18-25 years: results from a randomized controlled trial | Yes | Yes | Yes | Extracted | - | - |

| Author | Year | Title | Incident Infection | 6-month persistent infection | 12-month persistent infection | CIN2+ | Off-type efficacy | Immuno-genicity |
|---|---|---|---|---|---|---|---|---|
| Zhu[49] | 2014 | Immunogenicity and safety of the HPV-16/18 AS04-adjuvanted vaccine in healthy Chinese girls and women aged 9 to 45 years | - | - | - | - | - | Extracted |
| Zhu[50] | 2017 | Efficacy, immunogenicity, and safety of the HPV-16/18 AS04-adjuvanted vaccine in Chinese women aged 18-25 years: event-triggered analysis of a randomized controlled trial | Extracted | Extracted | Extracted | Yes | Extracted | Extracted |
| Welby[51] | 2022 | Progression from human papillomavirus (HPV) infection to cervical lesion or clearance in women (18-25 years): Natural history study in the control arm subjects of AS04-HPV-16/18 vaccine efficacy study in China between 2008 and 2016 | - | - | - | - | - | - |
| Zhao[52] | 2023 | Safety of AS04-HPV-16/18 vaccine in Chinese women aged 26 years and older and long-term protective effect in women vaccinated at age 18–25 years: A 10-year follow-up study | - | - | - | - | - | - |
| Wen[53] | 2025 | Ten-year evaluation of HPV vaccine influence on non-vaccine-type-related cervical infections and precancers in Chinese females: follow-up from a randomized control trial | - | - | - | - | - | - |

| Author | Year | Title | Incident Infection | 6-month persistent infection | 12-month persistent infection | CIN2+ | Off-type efficacy | Immuno-genicity |
|---|---|---|---|---|---|---|---|---|
| *China III (NCT00834106)* | | | | | | | | |
| Wen[54] | 2024 | The 13-year long-term follow-up on the effectiveness and immunogenicity of the quadrivalent human papillomavirus vaccine in Chinese females vaccinated at 20-45 years of age | - | - | - | Extracted | - | Extracted |
| Zhao[55] | 2023 | The eight-year long-term follow-up on the effectiveness of the quadrivalent human papillomavirus vaccine in Chinese women 20-45 years of age | - | - | - | - | - | - |
| *Future I (NCT00092521) & Future II (NCT00092534)*[†] | | | | | | | | |
| Ault[56] | 2007 | Effect of prophylactic human papillomavirus L1 virus-like-particle vaccine on risk of cervical intraepithelial neoplasia grade 2, grade 3, and adenocarcinoma in situ: a combined analysis of four randomised clinical trials | - | - | - | Pooled | - | - |

| Author | Year | Title | Incident Infection | 6-month persistent infection | 12-month persistent infection | CIN2+ | Off-type efficacy | Immuno-genicity |
|---|---|---|---|---|---|---|---|---|
| Perez[57] | 2008 | Safety, immunogenicity, and efficacy of quadrivalent human papillomavirus (types 6, 11, 16, 18) L1 virus-like-particle vaccine in Latin American women | - | - | - | - | - | - |
| Garland[58] | 2007 | Quadrivalent Vaccine against Human Papillomavirus to Prevent Anogenital Diseases | - | - | - | Extracted | - | - |
| Wheeler[59] | 2008 | Safety and immunogenicity of co-administered quadrivalent human papillomavirus (HPV)-6/11/16/18 L1 virus-like particle (VLP) and hepatitis B (HBV) vaccines | - | - | - | - | - | Extracted |
| FUTURE II[60] | 2007 | Prophylactic Efficacy of a Quadrivalent Human Papillomavirus (HPV) Vaccine in Women with Virological Evidence of HPV Infection | - | - | - | Pooled | - | - |
| Olsson[61] | 2009 | Evaluation of quadrivalent HPV 6/11/16/18 vaccine efficacy against cervical and anogenital disease in subjects with serological evidence of prior vaccine type HPV infection | - | - | - | Yes | - | - |

| Author | Year | Title | Incident Infection | 6-month persistent infection | 12-month persistent infection | CIN2+ | Off-type efficacy | Immuno-genicity |
|---|---|---|---|---|---|---|---|---|
| Kjaer[62] | 2009 | A pooled analysis of continued prophylactic efficacy of quadrivalent human papillomavirus (Types 6/11/16/18) vaccine against high-grade cervical and external genital lesions | - | - | - | - | Pooled | - |
| Brown[63] | 2009 | The impact of quadrivalent human papillomavirus (HPV; types 6, 11, 16, and 18) L1 virus-like particle vaccine on infection and disease due to oncogenic nonvaccine HPV types in generally HPV-naive women aged 16-26 years | | - | - | - | Pooled | - |
| Dillner[64] | 2010 | Four-year efficacy of prophylactic human papillomavirus quadrivalent vaccine against low grade cervical, vulvar, and vaginal intraepithelial neoplasia and anogenital warts: randomised controlled trial | - | - | - | Pooled | - | - |
| Munoz[65] | 2010 | Impact of human papillomavirus (HPV)-6/11/16/18 vaccine on all HPV-associated genital diseases in young women | - | - | - | Pooled | - | - |
| Haupt[66] | 2011 | Impact of an HPV6/11/16/18 L1 virus-like particle vaccine on progression to cervical intraepithelial neoplasia in seropositive women with HPV16/18 infection | - | - | - | - | - | - |

| Author | Year | Title | Incident Infection | 6-month persistent infection | 12-month persistent infection | CIN2+ | Off-type efficacy | Immuno-genicity |
|---|---|---|---|---|---|---|---|---|
| Joura[67] | 2012 | Effect of the human papillomavirus (HPV) quadrivalent vaccine in a subgroup of women with cervical and vulvar disease: retrospective pooled analysis of trial data | - | - | - | - | - | - |
| FUTURE II[68] | 2007 | Quadrivalent Vaccine against Human Papillomavirus to Prevent High-Grade Cervical Lesions | - | - | - | Extracted | - | - |
| Kjaer[69] | 2020 | Final analysis of a 14-year long-term follow-up study of the effectiveness and immunogenicity of the quadrivalent human papillomavirus vaccine in women from four Nordic countries | - | - | - | - | - | Extracted |
| Kjaer[70] | 2017 | A 12-Year Follow-up on the Long-Term Effectiveness of the Quadrivalent Human Papillomavirus Vaccine in 4 Nordic Countries | - | - | - | - | - | - |
| Lehtinen[71] | 2024 | Head-to-head comparison of two human papillomavirus vaccines for efficacy against cervical intraepithelial neoplasia grade 3 and adenocarcinoma in situ-population-based follow-up of two cluster-randomized trials | - | - | - | - | - | - |

| **Author** | **Year** | **Title** | **Incident Infection** | **6-month persistent infection** | **12-month persistent infection** | **CIN2+** | **Off-type efficacy** | **Immuno-genicity** |
|---|---|---|---|---|---|---|---|---|
| Joura[72] | 2026 | Effect of Prior 9-Valent Human Papillomavirus Vaccination on Subsequent Lower Genital Tract Dysplasia After Cervical Excisional Surgery | - | - | - | - | - | - |
| *Future III (NCT00090220)* | | | | | | | | |
| Castellsague[73] | 2011 | End-of-study safety, immunogenicity, and efficacy of quadrivalent HPV (types 6, 11, 16, 18) recombinant vaccine in adult women 24-45 years of age | - | Extracted | - | - | - | - |
| Luna[10] | 2013 | Long-term follow-up observation of the safety, immunogenicity, and effectiveness of Gardasil in adult women | - | - | - | Extracted | - | - |
| Maldonado[74] | 2022 | Effectiveness, immunogenicity, and safety of the quadrivalent HPV vaccine in women and men aged 27–45 years | - | - | - | - | - | Extracted |

| Author | Year | Title | Incident Infection | 6-month persistent infection | 12-month persistent infection | CIN2+ | Off-type efficacy | Immuno-genicity |
|---|---|---|---|---|---|---|---|---|
| Munoz[12] | 2009 | Safety, immunogenicity, and efficacy of quadrivalent human papillomavirus (types 6, 11, 16, 18) recombinant vaccine in women aged 24-45 years: a randomised, double-blind trial | - | Extracted | - | - | - | Yes |
| <u>*KenSHE*</u> | | | | | | | | |
| Barnabas[1] | 2022 | Efficacy of single-dose HPV vaccination among young African women | - | - | - | - | - | - |
| Barnabas[75] | 2023 | Durability of single-dose HPV vaccination in young Kenyan women: randomized controlled trial 3-year results | - | Extracted | - | - | - | - |
| Baisley[76] | 2024 | Comparing one dose of HPV vaccine in girls aged 9-14 years in Tanzania (DoRIS) with one dose in young women aged 15-20 years in Kenya (KEN SHE): an immunobridging analysis of randomised controlled trials | - | - | - | - | - | Extracted |

| Author | Year | Title | Incident Infection | 6-month persistent infection | 12-month persistent infection | CIN2+ | Off-type efficacy | Immuno-genicity |
|---|---|---|---|---|---|---|---|---|
| *Japan I*[‡] | | | | | | | | |
| Konno[77] | 2010 | Efficacy of human papillomavirus 16/18 AS04-adjuvanted vaccine in Japanese women aged 20 to 25 years: interim analysis of a phase 2 double-blind, randomized, controlled trial | Yes | Yes | - | - | - | Yes |
| Konno[78] | 2010 | Efficacy of human papillomavirus type 16/18 AS04-adjuvanted vaccine in Japanese women aged 20 to 25 years: final analysis of a phase 2 double-blind, randomized controlled trial | Yes | Extracted | Yes | Yes | Yes | Yes |
| Konno[79] | 2014 | Efficacy of the human papillomavirus (HPV)-16/18 AS04-adjuvanted vaccine against cervical intraepithelial neoplasia and cervical infection in young Japanese women | Extracted | - | Extracted | Extracted | Extracted | Extracted |
| *Japan II* | | | | | | | | |

| Author | Year | Title | Incident Infection | 6-month persistent infection | 12-month persistent infection | CIN2+ | Off-type efficacy | Immuno-genicity |
|---|---|---|---|---|---|---|---|---|
| Yoshikawa[80] | 2013 | Efficacy of quadrivalent human papillomavirus (types 6, 11, 16 and 18) vaccine (GARDASIL) in Japanese women aged 18-26 years | - | Extracted | - | - | - | Extracted |
| *Monovalent (NCT00365378)* | | | | | | | | |
| Koutsky[81] | 2002 | A controlled trial of a human papillomavirus type 16 vaccine | Extracted | Extracted | - | - | - | Yes |
| Mao[82] | 2006 | Efficacy of human papillomavirus-16 vaccine to prevent cervical intraepithelial neoplasia: a randomized controlled trial | - | Yes | - | Extracted | - | - |
| Rowhani-Rahbar[83] | 2009 | Longer term efficacy of a prophylactic monovalent human papillomavirus type 16 vaccine | Extracted | - | - | Extracted | - | Extracted |

| Author | Year | Title | Incident Infection | 6-month persistent infection | 12-month persistent infection | CIN2+ | Off-type efficacy | Immuno-genicity |
|---|---|---|---|---|---|---|---|---|
| *PATRICIA (NCT00122681)* | | | | | | | | |
| Garland[84] | 2016 | Prior human papillomavirus-16/18 AS04-adjuvanted vaccination prevents recurrent high grade cervical intraepithelial neoplasia after definitive surgical therapy: Post-hoc analysis from a randomized controlled trial | - | - | - | - | - | - |
| Lehtinen[85] | 2012 | Overall efficacy of HPV-16/18 AS04-adjuvanted vaccine against grade 3 or greater cervical intraepithelial neoplasia: 4-year end-of-study analysis of the randomised, double-blind PATRICIA trial | | | | | | Extracted |
| Paavonen[86] | 2007 | Efficacy of a prophylactic adjuvanted bivalent L1 virus-like-particle vaccine against infection with human papillomavirus types 16 and 18 in young women: an interim analysis of a phase III double-blind, randomised controlled trial | - | Yes | Yes | Yes | Yes | - |
| Paavonen[87] | 2009 | Efficacy of human papillomavirus (HPV)-16/18 AS04-adjuvanted vaccine against cervical infection and precancer caused by oncogenic HPV types (PATRICIA): final analysis of a double-blind, randomised study in young women | - | Yes | Yes | Yes | Yes | - |

| Author | Year | Title | Incident Infection | 6-month persistent infection | 12-month persistent infection | CIN2+ | Off-type efficacy | Immuno-genicity |
|---|---|---|---|---|---|---|---|---|
| Szarewski[88] | 2012 | Efficacy of the human papillomavirus (HPV)-16/18 AS04-adjuvanted vaccine in women aged 15-25 years with and without serological evidence of previous exposure to HPV-16/18 | Extracted | Extracted | Extracted | Extracted | - | - |
| Wheeler[89] | 2012 | Cross-protective efficacy of HPV-16/18 AS04-adjuvanted vaccine against cervical infection and precancer caused by non-vaccine oncogenic HPV types: 4-year end-of-study analysis of the randomised, double-blind PATRICIA trial | - | - | - | - | Extracted | - |
| Lehtinen[90] | 2017 | Ten-year follow-up of human papillomavirus vaccine efficacy against the most stringent cervical neoplasia end-point—registry-based follow-up of three cohorts from randomized trials | - | - | - | - | - | - |
| *Quadrivalent Phase II (NCT00365716)* | | | | | | | | |
| Villa[91] | 2005 | Prophylactic quadrivalent human papillomavirus (types 6, 11, 16, and 18) L1 virus-like particle vaccine in young women: a randomised double-blind placebo-controlled multicentre phase II efficacy trial | - | Yes | - | - | - | Yes |

| Author | Year | Title | Incident Infection | 6-month persistent infection | 12-month persistent infection | CIN2+ | Off-type efficacy | Immuno-genicity |
|---|---|---|---|---|---|---|---|---|
| Villa[92] | 2006 | Immunologic responses following administration of a vaccine targeting human papillomavirus Types 6, 11, 16, and 18 | - | - | - | - | - | Yes |
| Villa[93] | 2006 | High sustained efficacy of a prophylactic quadrivalent human papillomavirus types 6/11/16/18 L1 virus-like particle vaccine through 5 years of follow-up | - | Extracted | - | - | - | Extracted |
| *VIVIANE (NCT00294047)* | | | | | | | | |
| Skinner[94] | 2014 | Efficacy, safety, and immunogenicity of the human papillomavirus 16/18 AS04-adjuvanted vaccine in women older than 25 years: 4-year interim follow-up of the phase 3, double-blind, randomised controlled VIVIANE study | - | - | - | - | - | Extracted |
| Wheeler[95] | 2016 | Efficacy, safety, and immunogenicity of the human papillomavirus 16/18 AS04-adjuvanted vaccine in women older than 25 years: 7-year follow-up of the phase 3, double-blind, randomised controlled VIVIANE study | - | Extracted | - | Extracted | Extracted | - |

| Author | Year | Title | Incident Infection | 6-month persistent infection | 12-month persistent infection | CIN2+ | Off-type efficacy | Immuno-genicity |
|---|---|---|---|---|---|---|---|---|
| *<u>Quadrivalent in Men (NCT00090285)*</u> | | | | | | | | |
| Giuliano[96] | 2011 | Efficacy of Quadrivalent HPV Vaccine against HPV Infection and Disease in Males | Extracted | Extracted | - | - | - | - |
| Goldstone[97] | 2022 | Efficacy, immunogenicity, and safety of a quadrivalent HPV vaccine in men: results of an open-label, long-term extension of a randomised, placebo-controlled, phase 3 trial | - | - | - | - | - | - |
| Hillman[98] | 2012 | Immunogenicity of the quadrivalent human papillomavirus (type 6/11/16/18) vaccine in males 16 to 26 years old | - | - | - | - | - | Extracted |
| Palefsky[99] | 2011 | HPV Vaccine against Anal HPV Infection and Anal Intraepithelial Neoplasia | Extracted | Extracted | - | - | - | - |

| Author | Year | Title | Incident Infection | 6-month persistent infection | 12-month persistent infection | CIN2+ | Off-type efficacy | Immuno-genicity |
|---|---|---|---|---|---|---|---|---|
| Palefsky[100] | 2026 | Anal Human Papillomavirus Infection Progression to Disease Among Men Who Have Sex With Men | - | - | - | - | - | - |
| *uLACNet (NCT04255849)** | | | | | | | | |
| Giuliano[101] | 2025 | Design of a multicenter, randomized, double-blinded, placebo-controlled phase III trial evaluating the 9-valent human papillomavirus (HPV) vaccine to prevent persistent oral HPV infection in men living with human immunodeficiency virus: uLACNet trial 201 | - | - | - | - | - | - |

**Table S6.** A summary of the papers identified relating to the 17 RCTs identified that met out inclusion/exclusion criteria. † In the FUTURE I and FUTURE II trials, immunogenicity data was only identified for the FUTURE I study, though the FUTURE II protocol identified immunogenicity as an endpoint. In addition, multiple studies pooled participants from both trials to evaluate clinical efficacy and since there was no pooled immunogenicity data, this pooled efficacy estimates were not extracted and used. ‡ Japan I: The off-type efficacy data from this trial was extracted but not included in the analysis as only aggregated estimates of the off-type efficacy across multiple off-types were reported. *The RCTs in men only was excluded from our primary analyses. The extracted data is available as part of the published code along with details from each clinical trial.

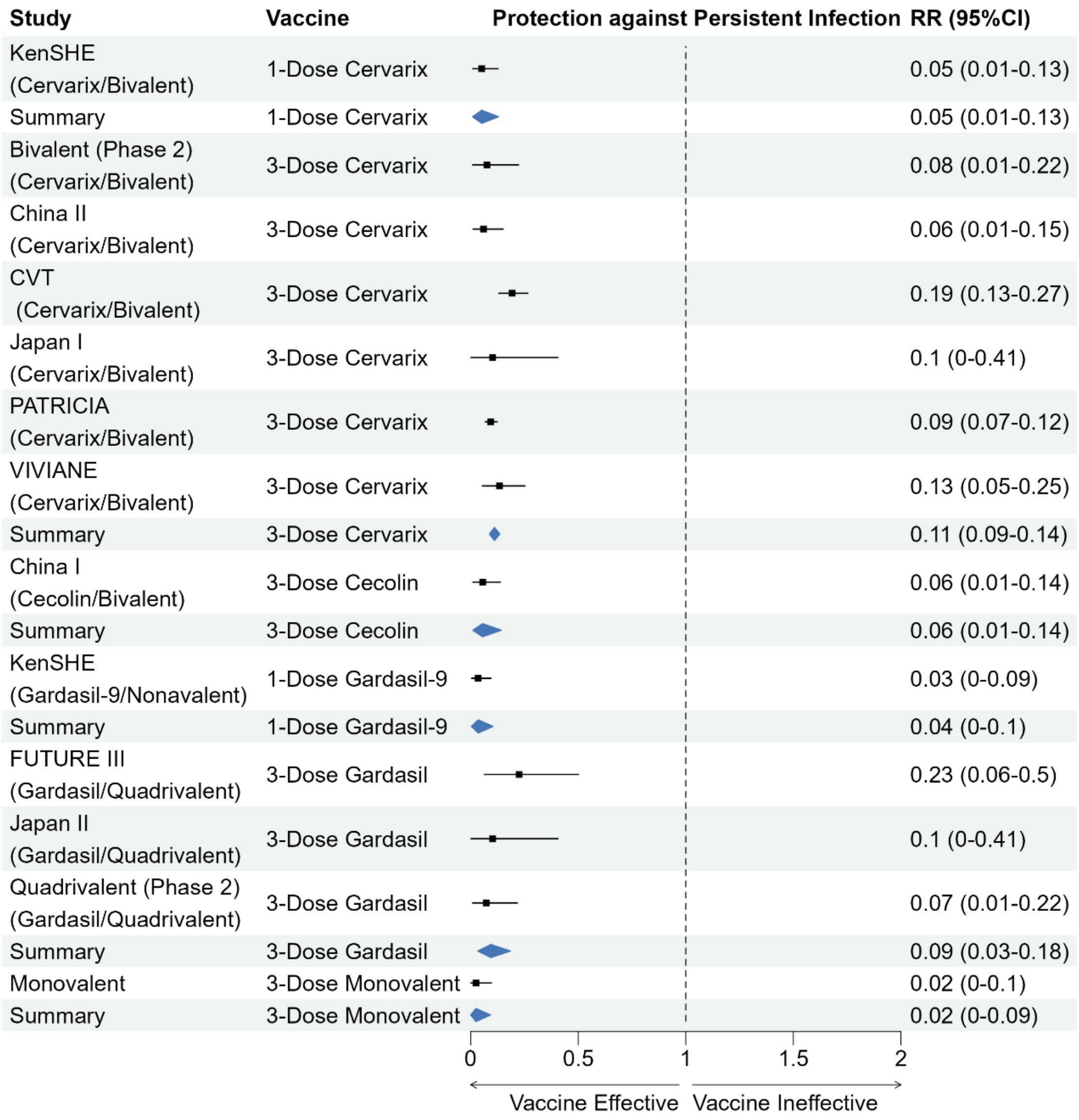


**Figure S2.** Forest plot of the vaccine efficacy against 6-month persistent infection with HPV 16

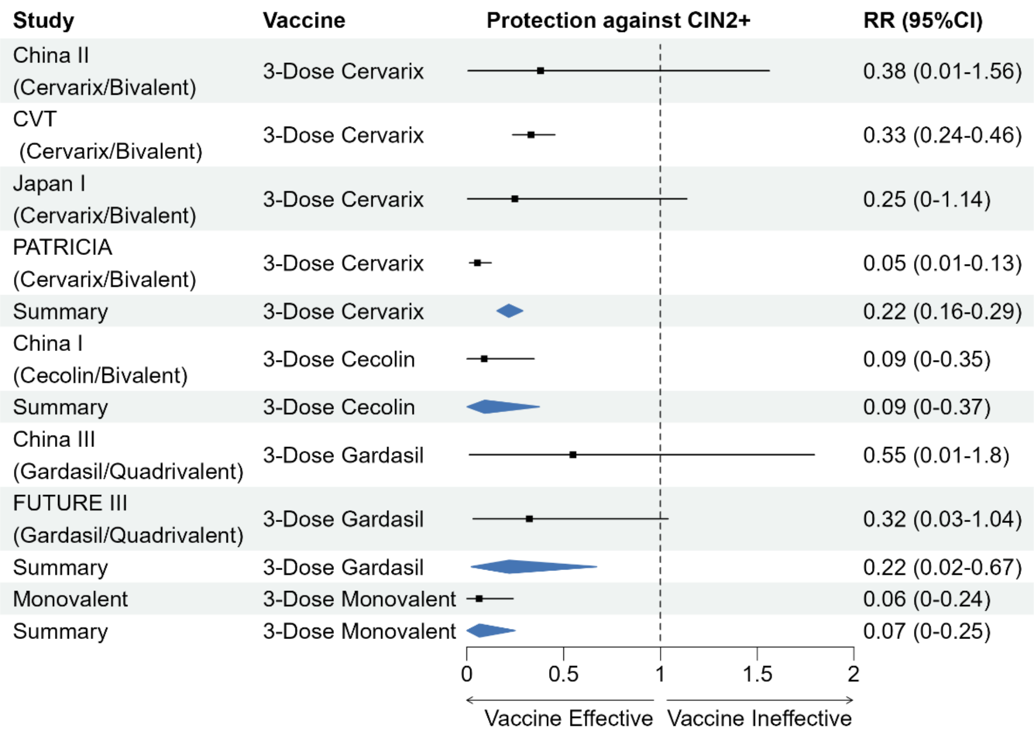


**Figure S3.** Forest plot of the vaccine efficacy against CIN2+ associated with HPV16.

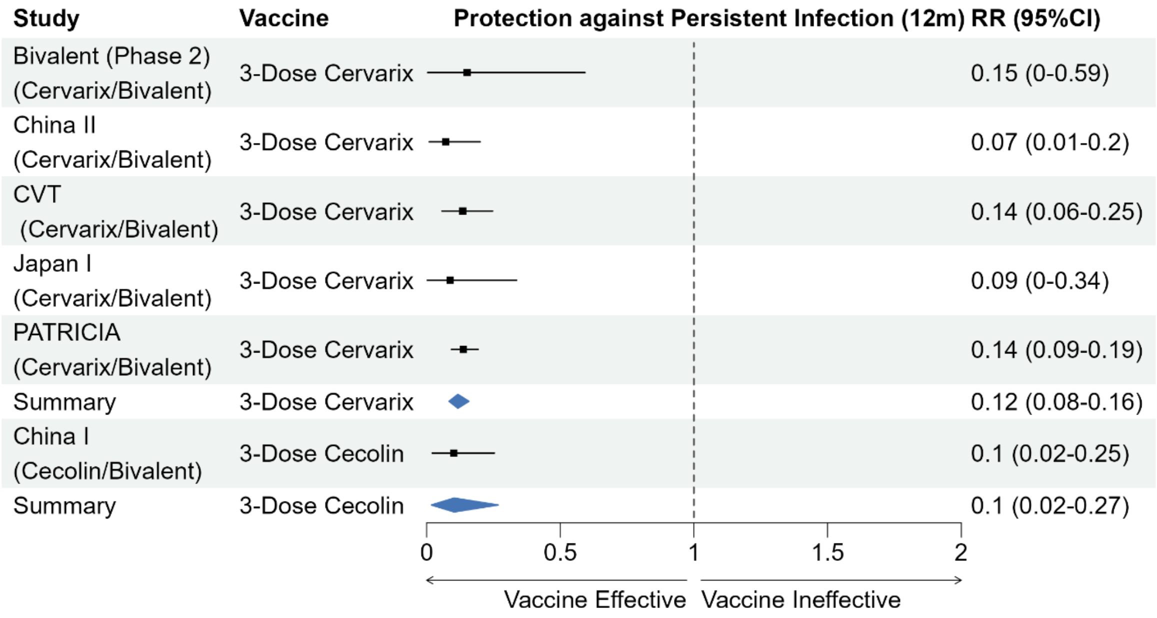


**Figure S4.** Forest plot of the vaccine efficacy against 12-month persistent infection with HPV16.

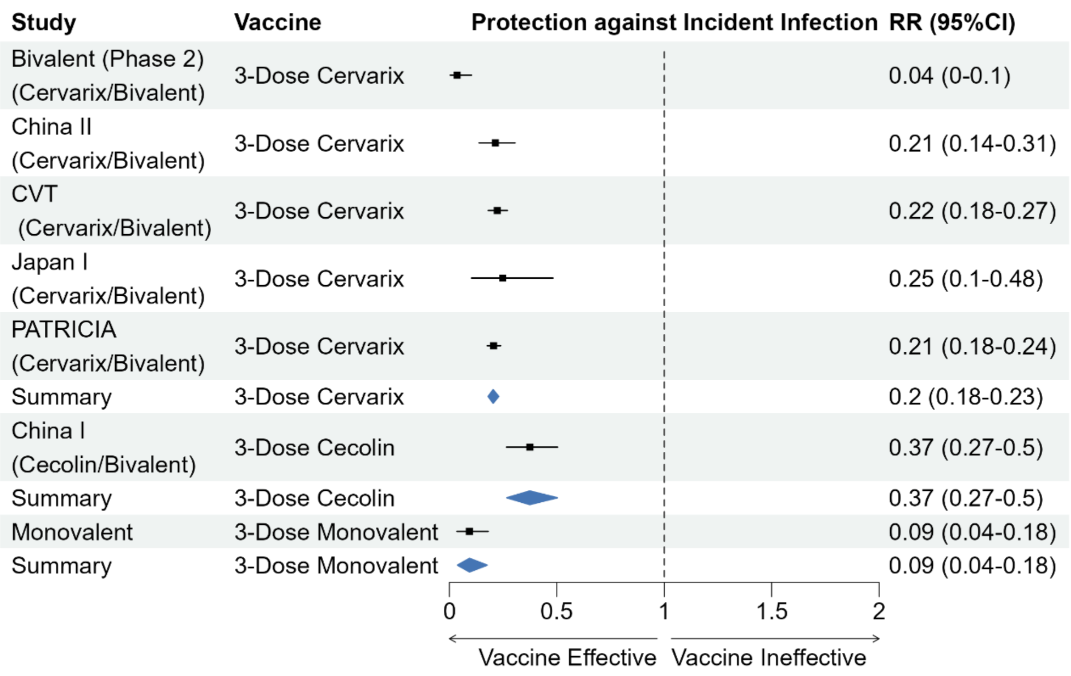


**Figure S5.** Forest plot of the vaccine efficacy against incident infection with HPV16.

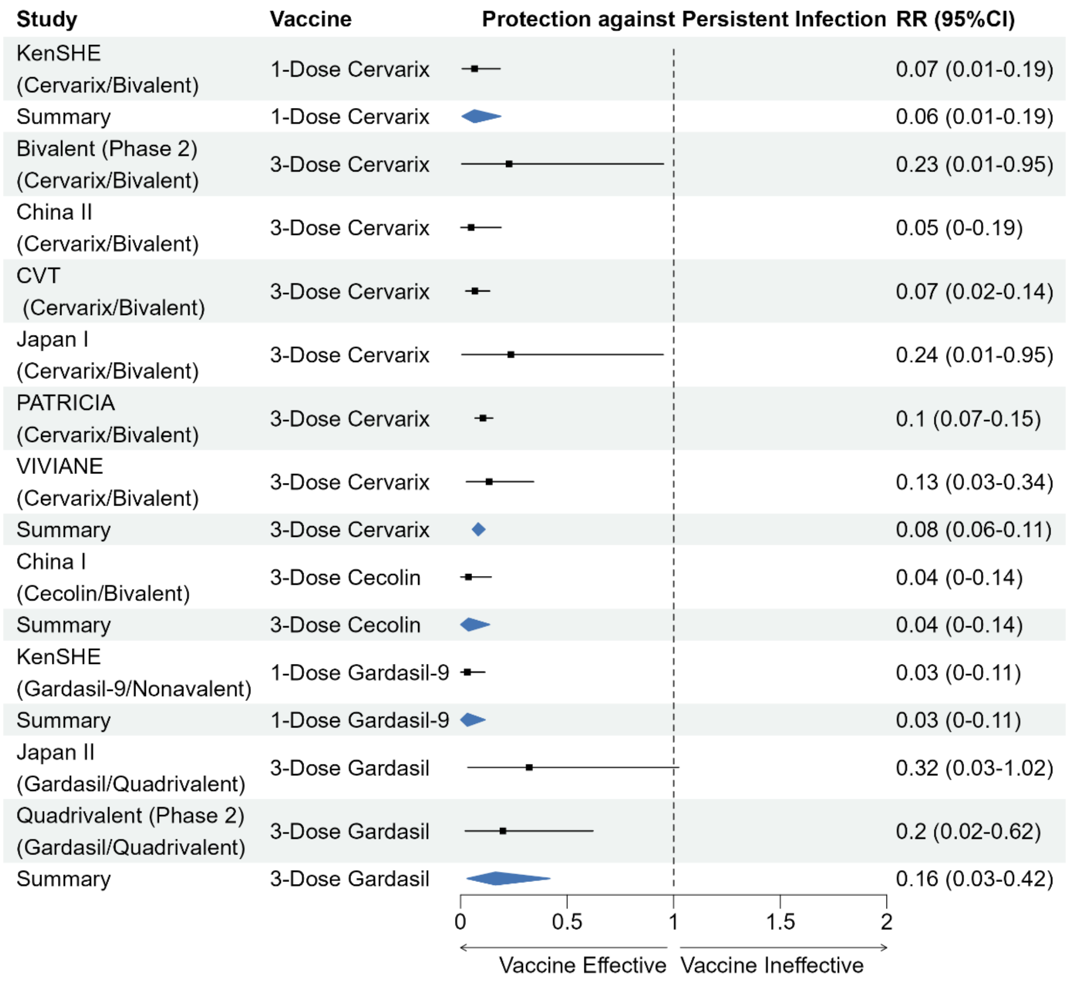


**Figure S6.** Forest plot of the vaccine efficacy against 6-month persistent infection with HPV18.

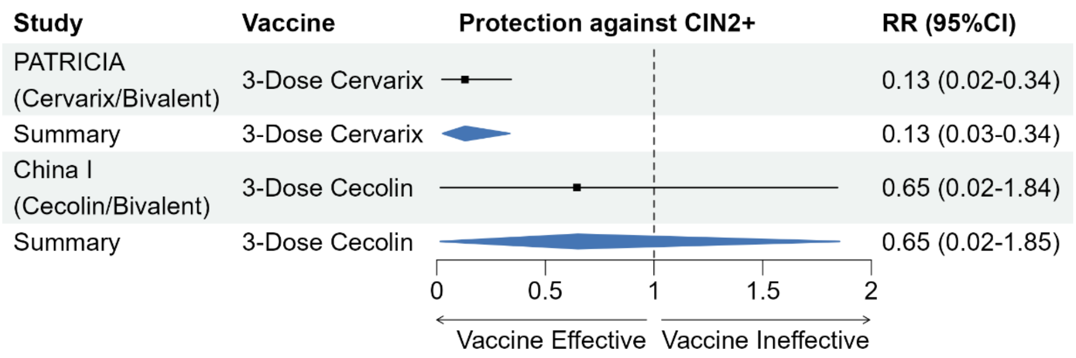


**Figure S7.** Forest plot of the vaccine efficacy against CIN2+ associated with HPV18.

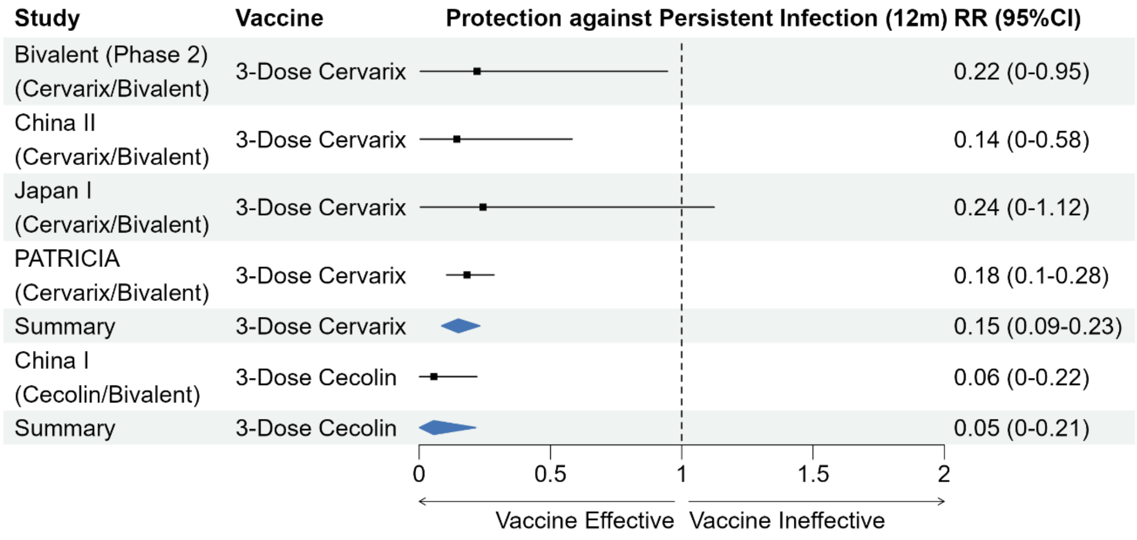


**Figure S8.** Forest plot of the vaccine efficacy against 12-month persistent infection with HPV18.

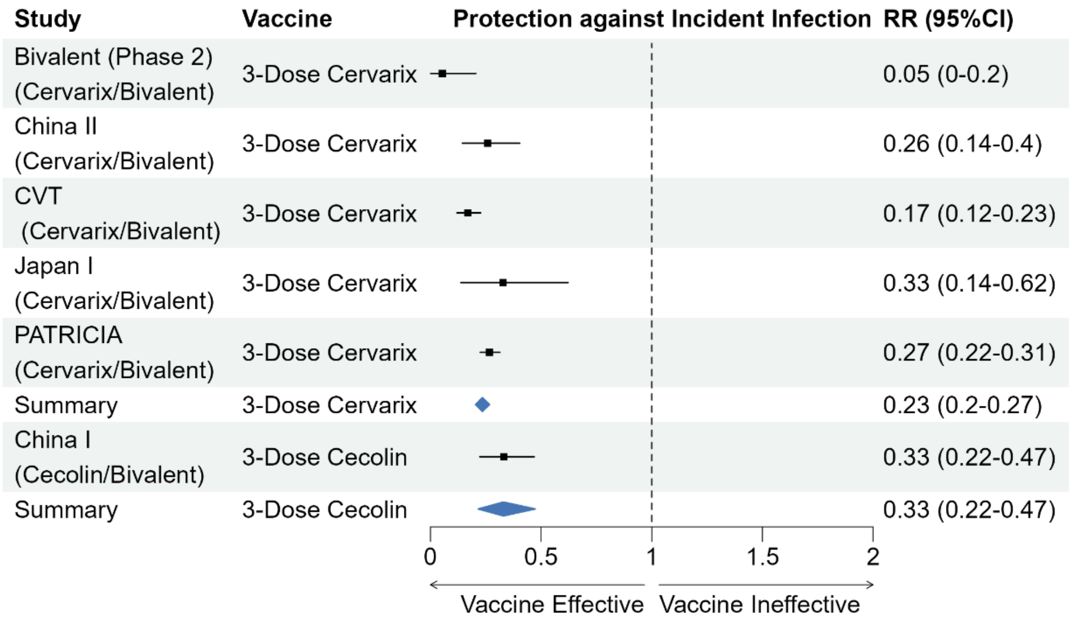


**Figure S9.** Forest plot of the vaccine efficacy against incident infection with HPV18.

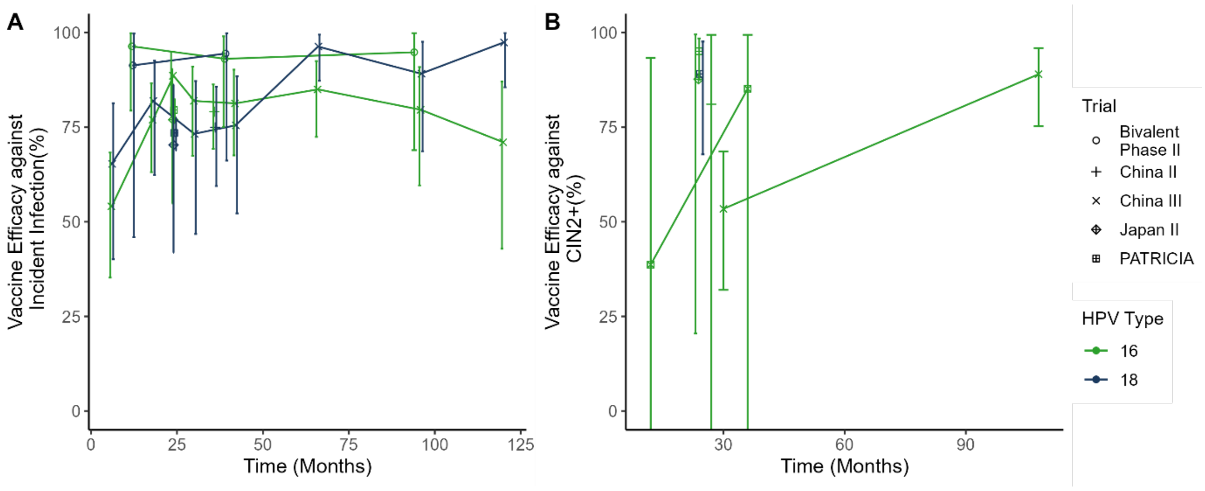


**Figure S10.** Vaccine efficacy over time (in months since first dose) measured in each of the trials (shapes) for the HPV types 16 and 18 (colours) against A) Incident infection and B) CIN2+.

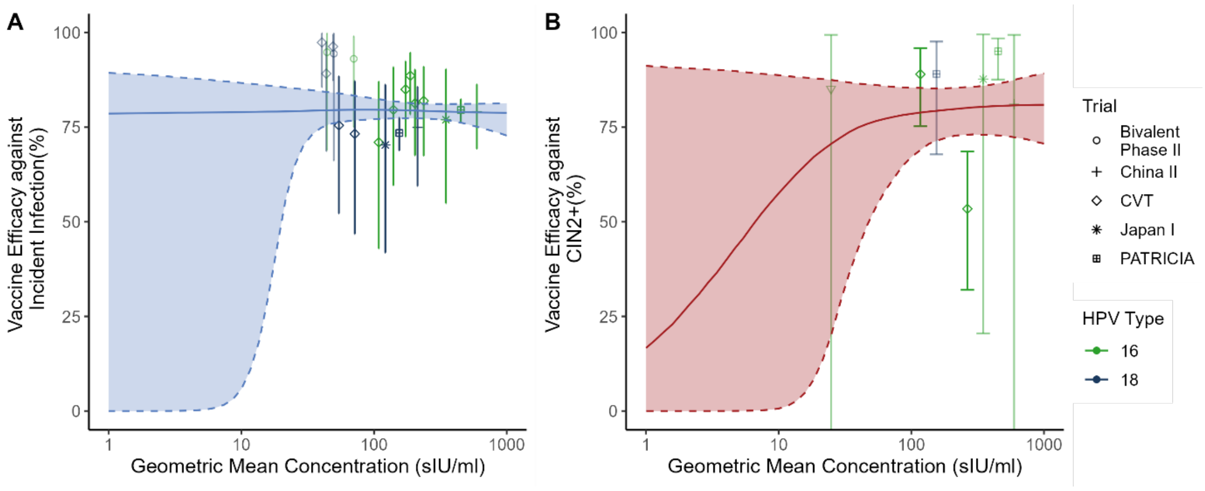


**Figure S11.** Testing for an association between antibody binding concentrations and vaccine efficacy against A) incident infection and B) CIN2+. The data points indicate observed vaccine efficacy and antibody binding concentrations from the available trials included in our analysis (shape) and against different HPV types (targeted by the vaccines, different types indicated by different colours). Error bars indicating the corresponding 95% credible intervals for vaccine efficacy. The solid line indicated the fitted logistic model relating antibody binding concentrations to vaccine efficacy, with 95% credible intervals indicated by the shaded regions.

| HPV type | GMC to non-vaccine HPV type (IU/mL) | Average fold-difference (overtime) $GMC_{16}$ / $GMC_X$ | Trial | HPV16 GMC at 12 months in RCT (IU/mL) | Estimated 12-month GMC to non-vaccine HPV type in RCT (scaled IU/mL) |
|---|---|---|---|---|---|
| 31 | 26 | 94 | CVT (Cervarix – 3 dose) | 384 | 4.1 |
| 33 | 11 | 225 | | | 1.7 |
| 45 | 51 | 47 | | | 8.0 |
| 52 | 10 | 236 | | | 1.6 |
| 58 | 20 | 122 | | | 3.1 |

| | | | | | |
|---|---|---|---|---|---|
| 31 | 26 | 94 | China II (Cervarix – 3 dose) | 664 | 7.0 |
| 33 | 11 | 225 | | | 2.9 |
| 45 | 51 | 47 | | | 13.8 |
| 52 | 10 | 236 | | | 2.8 |
| 58 | 20 | 122 | | | 5.4 |
| 31 | 26 | 94 | PATRICIA (Cervarix – 3 dose) | 621 | 6.5 |
| 33 | 11 | 225 | | | 2.8 |
| 45 | 51 | 47 | | | 12.9 |
| 52 | 10 | 236 | | | 2.6 |
| 58 | 20 | 122 | | | 5.1 |
| 31 | 26 | 94 | VIVIANE (Cervarix – 3 dose) | 289 | 3.1 |
| 33 | 11 | 225 | | | 1.3 |
| 45 | 51 | 47 | | | 6.0 |
| 52 | 10 | 236 | | | 1.2 |
| 58 | 20 | 122 | | | 2.4 |
| 31 | 2.9 | 49 | KenSHE (Cervarix – 1 dose) | 25 | 0.5 |
| 33 | 1.6 | 86 | | | 0.3 |
| 45 | 6.8 | 21 | | | 1.1 |
| 52 | 2.8 | 51 | | | 0.5 |
| 58 | 2.9 | 49 | | | 0.5 |
| 31 | 17.4 | 0.7 | KenSHE (Gardasil-9 – 1 dose) | 9.5 | 13.3 |
| 33 | 18.5 | 0.7 | | | 14.1 |
| 45 | 7.1 | 1.7 | | | 5.4 |
| 52 | 22.36 | 0.6 | | | 17.1 |
| 58 | 74 | 0.2 | | | 56.6 |

**Table S7.** Calculation of the estimated antibody concentration at 12 months after vaccination to HPV types 31, 33, 45, 52 and 58. These estimates are calculated for four randomised controlled trials that reported data on vaccine efficacy against these oncogenic HPV. Antibody concentrations are calculated by dividing the HPV16 GMC within each study by the average fold-differences (overtime) for each type relative to the HPV16 GMC in the two immunogenicity identified studies reporting these data[13,14].

| **Outcome** | **Excluded Study** | **Slope, $k$** | **Maximum Efficacy ($M$ %)** | **Concentration giving 50% of maximum efficacy ($EC_{50}$) (scaled IU/mL)** |
|---|---|---|---|---|
| Persistent Infection | None | 5.9 (4.7,7.6) | 90.6 (88.9,92) | 3.5 (3.2,3.9) |
| | Bivalent (Phase II) | 5.9 (4.7,7.5) | 90.4 (88.7,91.9) | 3.5 (3.2,3.9) |
| | China II | 5.9 (4.7,7.6) | 90.5 (88.7,91.9) | 3.2 (2.9,3.6) |
| | CVT | 7.2 (5.6,9.9) | 91.6 (89.8,93.2) | 4 (3.6,4.3) |
| | Future III | 5.7 (4.6,7.3) | 90.6 (89,92) | 3.5 (3.2,3.9) |
| | Japan I | 5.9 (4.7,7.5) | 90.4 (88.7,91.9) | 3.5 (3.2,4) |
| | Japan II | 5.9 (4.7,7.6) | 90.5 (88.9,92.1) | 3.5 (3.2,3.9) |
| | KenSHE | 4.5 (3.4,6) | 90.3 (88.4,92) | 3.8 (3.3,4.3) |
| | PATRICIA | 5.3 (4.1,7) | 90.7 (88.4,92.7) | 2.9 (2.4,3.4) |
| | Quadrivalent (Phase II) | 5.8 (4.6,7.4) | 90.4 (88.6,91.8) | 3.5 (3.2,3.9) |
| | VIVIANE | 6 (4.8,7.8) | 90.6 (88.8,92.2) | 3.6 (3.2,4) |
| | None | 2.7 (2,3.7) | 79 (76.8,81.3) | 2.6 (2.1,3.3) |

| | | | | |
|---|---|---|---|---|
| Incident Infection | Bivalent (Phase 2) | 2.6 (1.8,3.6) | 78.6 (76.3,81.2) | 2.6 (2.1,3.4) |
| | China II | 4.6 (3.4,6.1) | 78.8 (76.7,80.7) | 1.7 (1.4,2) |
| | CVT | 3 (1.5,5.2) | 79 (76.1,86.4) | 6.2 (4.2,11.4) |
| | Japan I | 2.7 (2,3.7) | 79 (76.9,81.4) | 2.6 (2.1,3.3) |
| | PATRICIA | 2.7 (2,3.6) | 81.1 (77.8,84.5) | 2.7 (2.2,3.5) |
| CIN2+ | None | 3.6 (1.4,6.4) | 81.4 (75.2,87.5) | 2 (1.2,3.5) |
| | China II | 3.7 (1.6,6.5) | 81 (74.9,86.8) | 2 (1.1,3.5) |
| | CVT | 2.1 (1.1,4.6) | 95.8 (89.9,99.7) | 1.7 (0.6,3.3) |
| | FUTURE III | 3.6 (1.4,6.5) | 81.3 (74.9,87.4) | 2 (1.1,3.8) |
| | Japan I | 3.7 (1.6,6.4) | 80.8 (74.3,86.6) | 2 (1.1,3.4) |
| | PATRICIA | 5.6 (1.9,12.5) | 70.5 (58.9,79.8) | 16.3 (3.1,44.5) |

**Table S8.** Leave-one-out analysis for persistent infection, incident infection and CIN2+ correlate of protection analysis. Here we show the impact of leaving one RCT out of the model fitting when estimating the relationship between antibody binding titres and vaccine efficacy. That is, we re-fitted the data on each outcome (as in Figure 2) but with one RCT excluded in each case. We find that parameter estimates are robust to the exclusion of any one trial for persistent infection. We find that the $EC_{50}$ had some sensitivity to the CVT and China II studies for incident infection, as these were the only studies which included HPV types 31, 33, 45, 52 and 58. For CIN2+, the maximum efficacy has some sensitivity to the exclusion of the CVT and PATRICIA trials, and the $EC_{50}$ showed some sensitivity to the exclusion of the PATRICIA trial.

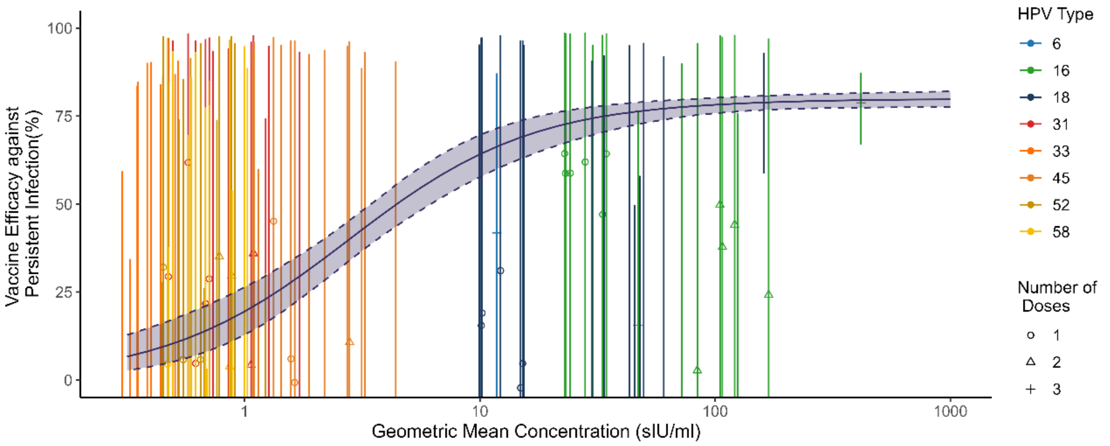


**Figure S12.** Comparing our correlate of protection (fitted in Figure 2C), to vaccine efficacy data from non-per-protocol estimates reported in some of the included randomised controlled trials. The fitted logistic model from Figure 2C in the main text is shown by the solid line with shaded region indicating the 95% credible interval. The data points are the vaccine efficacy estimates for non-per-protocol arms of the studies, including those who received a reduced number of doses (shape) or who received the full regimen but were seropositive at baseline (a cohort intended to be excluded from the trial). Data is grouped by the HPV type being included in the vaccine (green points) or cross-protective HPV types (orange points). Note that the credible intervals are large as many of the reduced dose arms contain a small population and incidence rates.

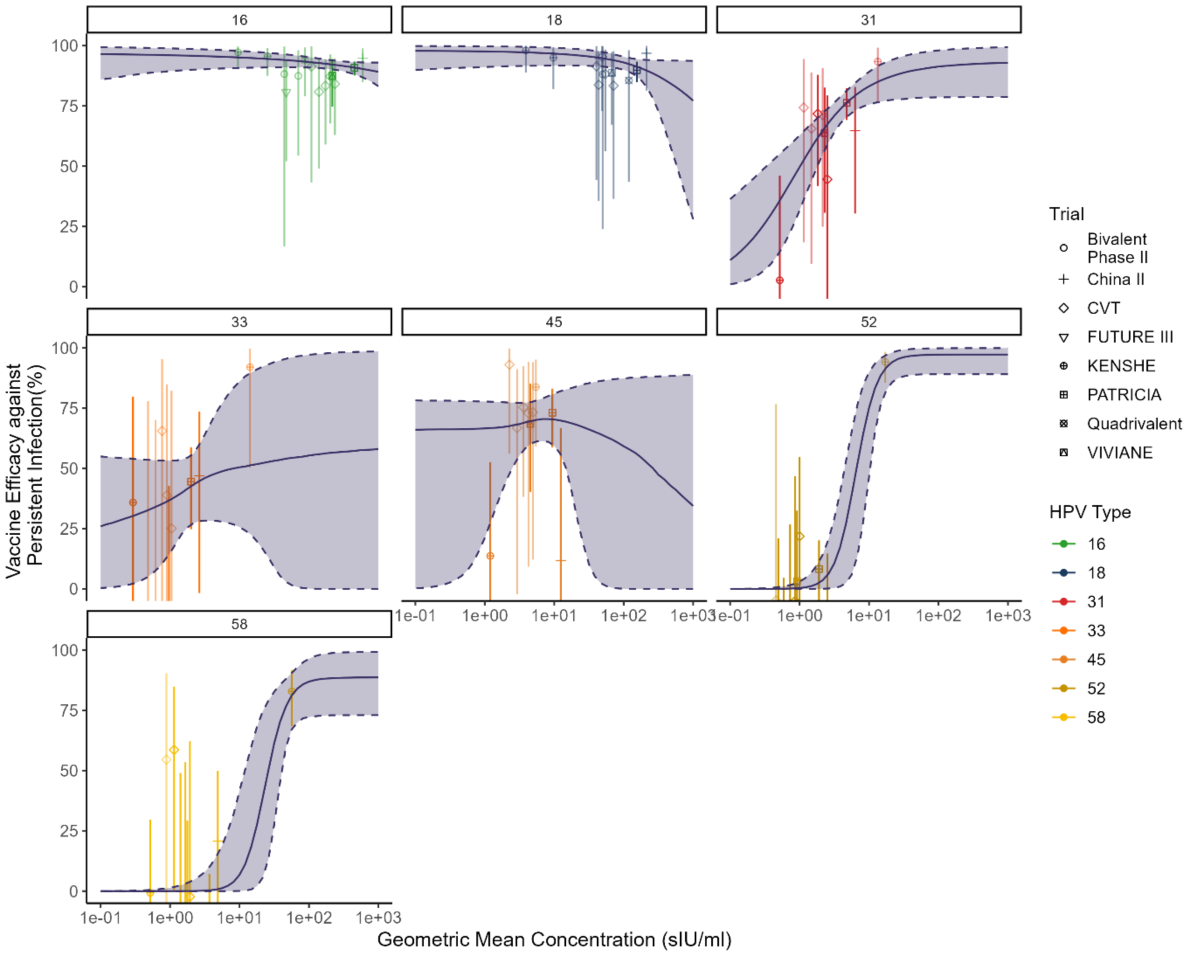


**Figure S13.** Comparison of the estimated logistic relation between antibody binding concentration and vaccine efficacy against persistent infection stratifying by HPV type (i.e. each type is fitted independently).

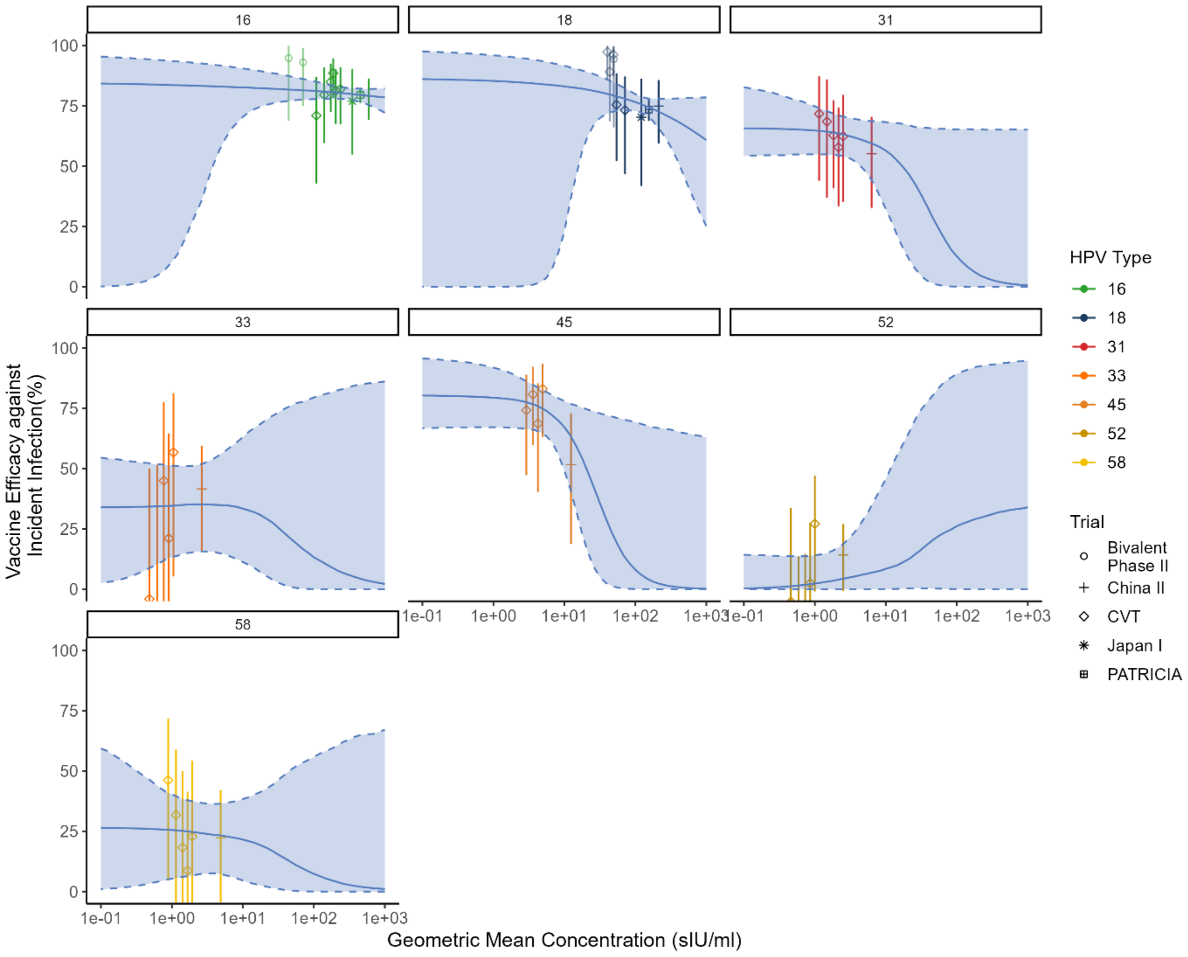


**Figure S14.** Comparison of the estimated logistic relation between antibody binding concentration and Vaccine efficacy against incident infection stratifying by HPV type (i.e. each type is fitted independently). Data is too limited to comment on any HPV type specific associations between binding and vaccine efficacy against incident infection. All data HPV type 31, 33, 45, 52 and 58 data is from either the CVT or China II study, both of which used the 3-dose Cervarix regimen.

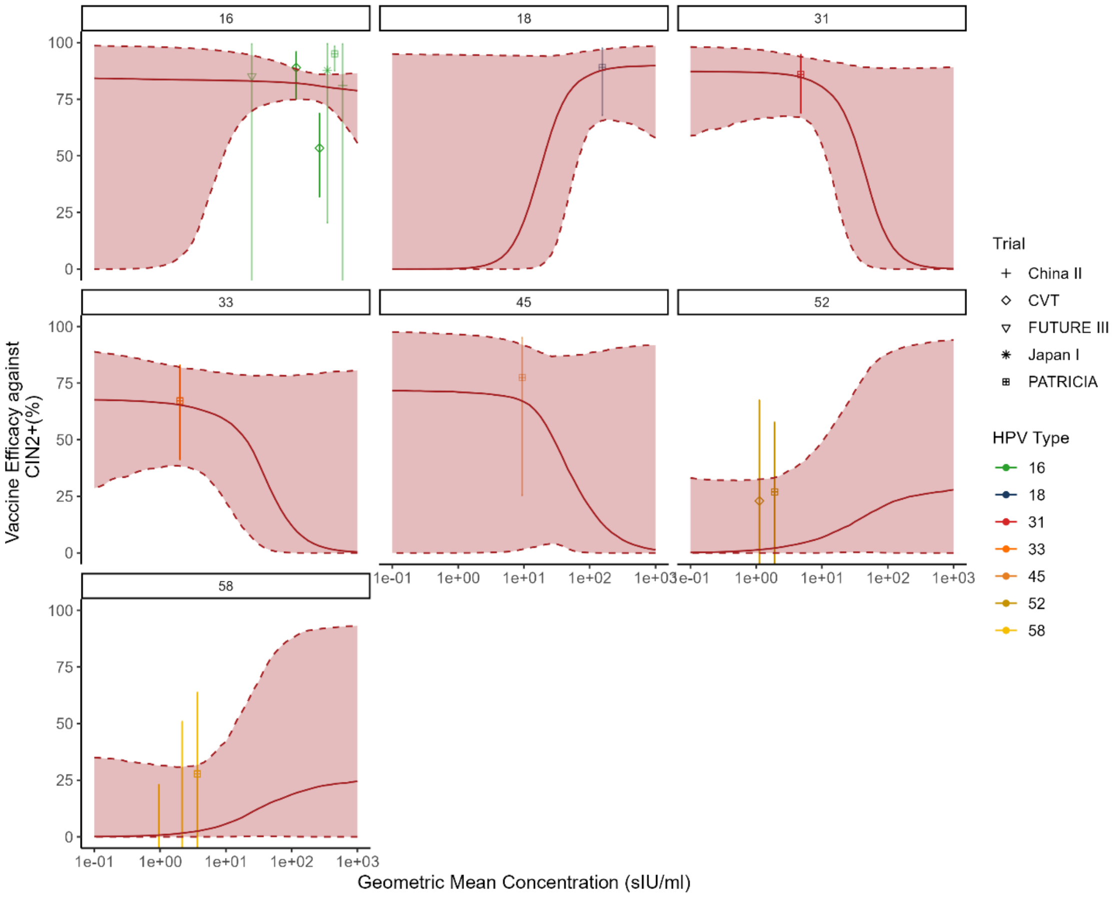


**Figure S15.** Comparison of the estimated logistic relation between antibody binding concentration and Vaccine efficacy against CIN2+ assessing each HPV type independently. Data is too limited to comment on any HPV type specific associations between binding and vaccine efficacy against CIN2+. All data HPV type 31, 33, 45, 52 and 58 data is from the CVT, which used the 3-dose Cervarix regimen.

| HPV Type | 50% Protective Concentration ($EC_{50}$, sIU/ml) |
|---|---|
| 6 | 1.3 (0.10,4.0) |
| 11 | 13.1(0.9,224) |
| 16 | 0.54 (0.047,1.9) |
| 18 | 0.28(0.023,1.1) |
| 31 | 1.38 (0.9,1.9) |
| 33 | 2.19 (1.4,3.8) |
| 45 | 2.21 (1.4,3.4) |
| 52 | 6.26 (4.4,9.4) |
| 58 | 16.8 (9.8,29.8) |

**Table S9.** Refitting our correlate model allowing the EC50 to differ between types. Here we show that there are significant differences between the EC50 for the different HPV types whilst using the same slope and maximum efficacy across all HPV types.

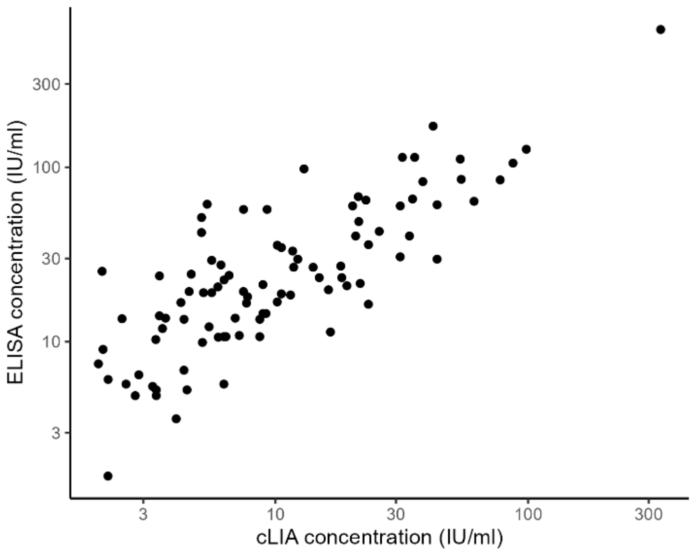


**Figure S16**. Comparison of the two main antibody binding assays used in our analysis, using data from Safaeian et al.[102] This study reported binding antibody concentrations against HPV16of a set of serum samples using the same ELISA and a competitive binding Luminex assay (cLIA) assays used in our analysis. Extracting this data, we found that these two assays were well correlated.

| **Outcome** | **Assay** | **Slope, *k*** | **Maximum Efficacy (*M* %)** | **Concentration giving 50% of maximum efficacy ($EC_{50}$)** |
|---|---|---|---|---|
| Persistent Infection | Combined analysis | 6.1 (4.8-8.0) | 91.4 (89.7-92.9) | 3.6 (3.3-4.0) |
| | ELISA | 5.4 (4.4-6.9) | 90.7 (88.9-92.2) | 3.5 (3.2-3.9) |
| | cLIA | 5.4 (4.4-6.9) | 90.7 (88.9-92.2) | 2.9 (0.6-6.1) |
| CIN2+ | Combined analysis | 3.7 (1.2- 6.5) | 81.5 (75.1-89.7) | 2.0 (1.1-3.7) |
| | ELISA | 3.5 (1.1-6.3) | 81.4 (75.3-89.8) | 2.0 (1.1-3.9) |
| | cLIA | 3.5 (1.1-6.3) | 81.4 (75.3-89.8) | 13.2 (0.3-154) |

**Table S10.** The estimated parameters when applying a fixed assay effect allowing the EC50 parameters to differ between the two different assays. Incidence data was all assessed on the ELISA assay.

| **Parameter** | **Antibody model estimates** | **Neutralisation model estimates** |
|---|---|---|
| Maximum efficacy (M) | 91.0% (89.3-92.5%) | 92.7% (86.2-97.1%) |
| Slope (k) | 4.9 (3.9-6.3) | 3.6 (2.0-6.2) |
| $EC_{50}$ (level at which 50% of maximum efficacy is achieved) | 24 (21-27) | 82 (47-144) |

**Table S11.** Comparison of the model parameters using neutralisation titres (from Mariz et. al[19]) as a correlate and using binding antibody concentrations as a correlate.

| Study or title | Country | Population | Vaccine | Study design | Study endpoints |
|---|---|---|---|---|---|
| **RCTs** | | | | | |
| FUTURE (group) | Australia | 20,583 women aged 16–26 years | Merck 4vHPV | RCT | VE against HPV infection. |
| KENSHE | Kenya | 2,250 sexually active females aged 15–20 y. | GSK 2vHPV, Merck 9vHPV | RCT | VE against HPV infection, humoral & cellular immunogenicity, cost-effectiveness. |
| CVT | Costa Rica | 1,000 females vaccinated aged 18–25 y | GSK 2vHPV | Long-term FU study of participants previously vaccinated with 1d v 2d v 3d through an RCT | Humoral immunogenicity |
| IARC India | India | 1,540 age-matched, & unvaccinated females | Merck 4vHPV | Observational cohort study of 1d v 2d v 3d, and v no vaccination (extended FU) | VE against HPV infection; humoral immunogenicity |
| DoRIS | Tanzania | 930 females aged 9–14 y | GSK 2vHPV, Merck 9vHPV | RCT of 1d v 2d v 3d | Humoral & cellular immunogenicity; cost-effectiveness; acceptability |
| **Cochrane reviews** | | | | | |
| Prophylactic vaccination against human papillomaviruses to prevent cervical cancer and its precursors (2018). | | | GSK 2vHPV, Merck 4vHPV | Systematic review & meta-analysis. | Efficacy and immunogenicity |
| Comparison of different human papillomavirus (HPV) vaccine types and dose schedules for prevention of HPV-related disease in | | | GSK 2vHPV, Merck 4vHPV Merck 9vHPV | Systematic review & meta-analysis. | Efficacy and immunogenicity |

| | | | | |
|---|---|---|---|---|
| females and males (2019). | | | | |
| Efficacy, effectiveness and immunogenicity of one dose of HPV vaccine compared with no vaccination, two doses, or three doses (2022) | | GSK 2vHPV, Merck 4vHPV | Systematic review & meta-analysis. | Efficacy and immunogenicity |

**Table S12.** Overview of scoping review identifying the characteristics of key trials and the Cochrane reviews

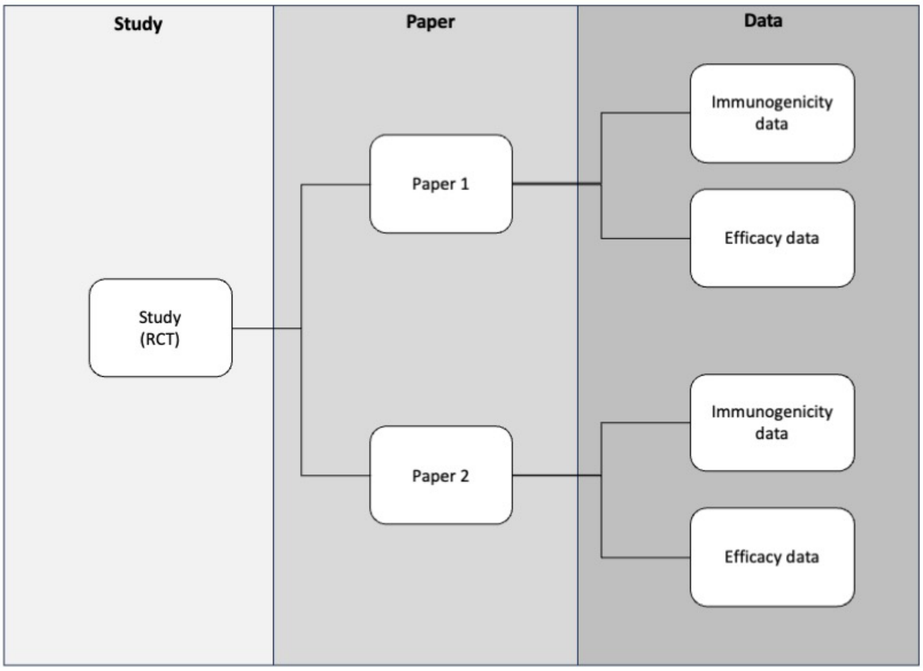


**Figure S17.** Outline of the extraction structure for the immunogenicity and efficacy data from RCTs.

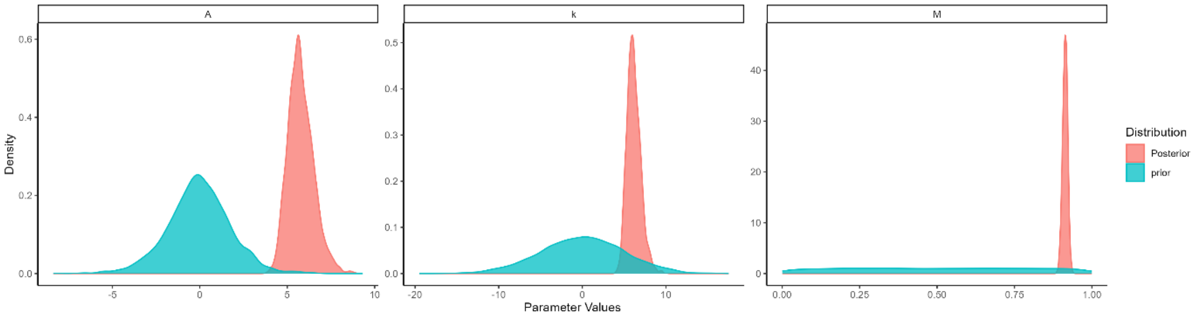


**Figure S18.** Comparison of the prior and posterior distributions using the model fit against persistent infection. (Left) Parameter A, (Middle) Slope Parameter, k, and (Right) Maximum efficacy, M.

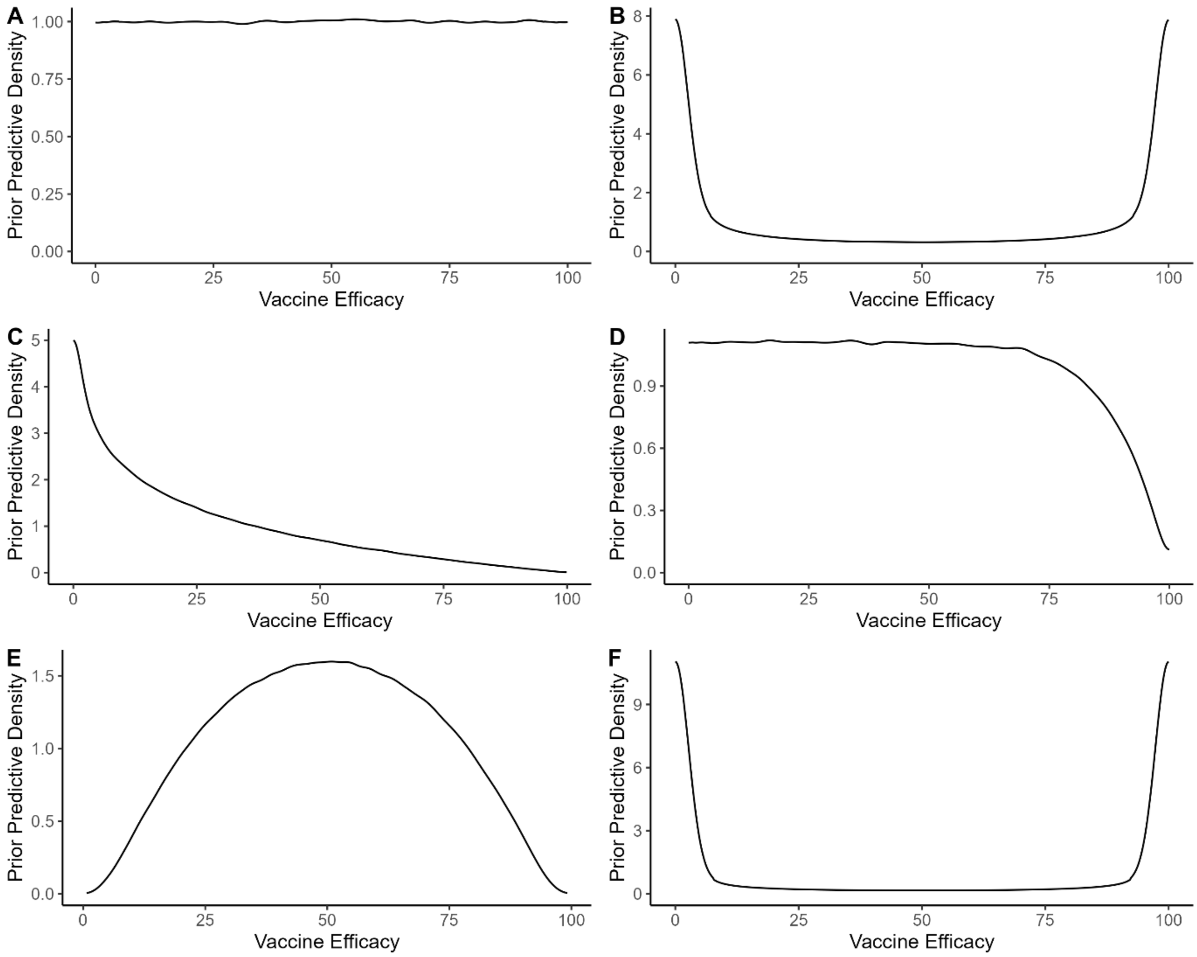


**Figure S19.** Prior predictive vaccine efficacy for different choices of prior distributions using (A-D) a fixed slope, k=0 (assuming no correlation). A) Logistic distribution for A, with the maximum fixed $M = 1$. B) Using a normal prior $A \sim N(0,1)$, with the maximum fixed, $M = 1$. C) Using a logistic prior for A and a uniform prior for M. D) Using a logistic prior for A and a Beta distribution for M, $M \sim Beta(10,1)$. The prior predictive vaccine efficacy at a GMC 10-fold above the EC50 concentration using E) a narrow prior, $k \sim N(0,1)$ or F) a diffuse prior $k \sim N(0,10)$.

| Outcome | Parameter | Estimates using different priors | | | |
|---|---|---|---|---|---|
| | | Uniform(0,1) | Beta (5,1) | Beta(10,1) | Beta (20,1) |
| Incident Infection | Maximum Efficacy, M | 79.9 (77.6,82.5) | 79.9 (77.7,82.4) | 80.0 (77.7,82.8) | 80.2 (77.8,83.3) |
| Persistent Infection | | 91.4 (89.7,93.0) | 91.5 (89.7,93.0) | 91.5 (89.9,92.9) | 91.6 (89.9,93.0) |
| CIN2+ | | 81.4 (75.3,87.1) | 81.9 (75.9,89.0) | 82.4 (76.4,89.2) | 83.7 (77.5,98.3) |
| Incident Infection | Slope, k | 2.5 (1.8-3.5) | 2.5 (1.8,3.5) | 2.5 (1.8,3.4) | 2.5 (1.7,3.5) |
| Persistent Infection | | 6.2 (4.9-8.3) | 6.2 (4.9,8.3) | 6.2 (4.9,8.1) | 6.2 (4.9,8.2) |
| CIN2+ | | 3.7 (1.6-6.5) | 3.7 (1.2,6.5) | 3.6 (1.2,6.5) | 3.4 (0.8,6.5) |

**Table S13**: Comparison of the posterior 95% credible intervals for the maximum efficacy, M, and $k$, using different prior distributions for $M$.

| Outcome | Parameter | Estimates of the parameter, A, for different priors | | | |
|---|---|---|---|---|---|
| | | Logistic(0,1) | Logistic(0,10) | Normal(0,1) | Normal(0,10) |
| Incident Infection | A | 2.7 (1.8,3.7) | 2.9 (2.0,4.0) | 2.4 (1.6,3.3) | 2.9 (2,4.1) |
| Persistent Infection | | 5.8 (4.7,7.6) | 6.5 (5,9.1) | 4.6 (3.9,5.5) | 6.4 (5,9.1) |
| CIN2+ | | 4.4 (1.9,7.5) | 7.2 (3.6,12.7) | 2.1 (0.8,3.7) | 7.1 (3.6,12.8) |
| Incident Infection | Slope, k | 2.5 (1.8-3.5) | 2.7 (1.9,3.8) | 2.3 (1.6,3.1) | 2.7 (2.0,3.8) |
| Persistent Infection | | 6.2 (4.9-8.3) | 6.9 (5.2,10) | 4.9 (4,5.9) | 6.9 (5.3,10.1) |
| CIN2+ | | 3.7 (1.6-6.5) | 6.1 (3.0,10.8) | 1.8 (0.8,3.2) | 6.0 (3.0,11) |

**Table S14**: Comparison of the posterior 95% credible intervals for the parameter, A and $k$ using different prior distributions for $A$.

| Outcome | Estimates of the slope parameter, k, for different priors | | | |
|---|---|---|---|---|
| | Normal(0,1) | Normal(0,5) | Normal(0,10) | Normal(0,20) |
| Incident Infection | 2.2 (1.6,3.0) | 2.5 (1.8,3.5) | 2.6 (1.8,3.5) | 2.6 (1.8,3.5) |
| Persistent Infection | 4.6 (3.8,5.5) | 6.2 (4.9,8.3) | 6.3 (5.0,8.5) | 6.3 (4.9,8.4) |
| CIN2+ | 1.7 (0.8,3.2) | 3.7 (1.8,6.5) | 3.9 (1.7,7.1) | 3.9 (1.7,7.1) |

**Table S15**: Comparison of the posterior 95% credible intervals for the slope parameter, k, using different prior distributions for $k$.

### References


1. Barnabas Ruanne, V., *et al.* Efficacy of Single-Dose Human Papillomavirus Vaccination among Young African Women. *NEJM Evidence* **1**, EVIDoa2100056 (2022).
2. Sankaranarayanan, R., *et al.* Can a single dose of human papillomavirus (HPV) vaccine prevent cervical cancer? Early findings from an Indian study. *Vaccine* **36**, 4783-4791 (2018).
3. Herrero, R., *et al.* Prevention of Persistent Human Papillomavirus Infection by an HPV16/18 Vaccine: A Community-Based Randomized Clinical Trial in Guanacaste, Costa Rica. *Cancer Discovery* **1**, 408-419 (2011).
4. Single-Dose HPV Vaccine Evaluation Consortium. Review of the current published evidence for single-dose HPV vaccination. (PATH, 2022).
5. Arbyn, M., Xu, L., Simoens, C. & Martin-Hirsch, P.P.L. Prophylactic vaccination against human papillomaviruses to prevent cervical cancer and its precursors. *Cochrane Database of Systematic Reviews* (2018).
6. Bergman, H., *et al.* Comparison of different human papillomavirus (HPV) vaccine types and dose schedules for prevention of HPV-related disease in females and males. *Cochrane Database of Systematic Reviews* (2019).
7. Henschke, N., *et al.* Efficacy, effectiveness and immunogenicity of one dose of HPV vaccine compared with no vaccination, two doses, or three doses. (Cochrane Response, 2022).
8. Naud, P.S., *et al.* Sustained efficacy, immunogenicity, and safety of the HPV-16/18 AS04-adjuvanted vaccine: final analysis of a long-term follow-up study up to 9.4 years post-vaccination. *Hum Vaccin Immunother* **10**, 2147-2162 (2014).
9. Beachler, D.C., *et al.* Multisite HPV16/18 Vaccine Efficacy Against Cervical, Anal, and Oral HPV Infection. *J Natl Cancer Inst* **108**(2016).
10. Luna, J., *et al.* Long-term follow-up observation of the safety, immunogenicity, and effectiveness of Gardasil in adult women. *PLoS One* **8**, e83431 (2013).
11. Zhao, F., *et al.* Long-term efficacy and immunopersistence of an -Escherichia coli-produced HPV-16/18 bivalent vaccine: an observational extension study following a randomised, double-blind Phase III clinical trial cohort. *The Lancet Regional Health – Western Pacific* **61**(2025).
12. Munoz, N., *et al.* Safety, immunogenicity, and efficacy of quadrivalent human papillomavirus (types 6, 11, 16, 18) recombinant vaccine in women aged 24-45 years: a randomised, double-blind trial. *Lancet* **373**, 1949-1957 (2009).
13. Pasmans, H., *et al.* Long-term HPV-specific immune response after one versus two and three doses of bivalent HPV vaccination in Dutch girls. *Vaccine* **37**, 7280-7288 (2019).
14. Kemp, T.J., *et al.* Immunogenicity of one and two doses of Gardasil®9 in Tanzanian girls in the DoRIS Trial. *npj Vaccines* **11**, 22 (2025).
15. Toh, Z.Q., *et al.* Sustained Antibody Responses 6 Years Following 1, 2, or 3 Doses of Quadrivalent Human Papillomavirus (HPV) Vaccine in Adolescent Fijian Girls, and Subsequent Responses to a Single Dose of Bivalent HPV Vaccine: A Prospective Cohort Study. *Clinical Infectious Diseases* **64**, 852-859 (2017).
16. Berry, M.T., *et al.* Predicting vaccine effectiveness for mpox. *Nat Commun* **15**, 3856 (2024).
17. Elias, K.M., *et al.* Viral clearance as a surrogate of clinical efficacy for COVID-19 therapies in outpatients: a systematic review and meta-analysis. *The Lancet Microbe* **5**, e459-e467 (2024).
18. Khoury, D.S., *et al.* Neutralizing antibody levels are highly predictive of immune protection from symptomatic SARS-CoV-2 infection. *Nat Med* **27**, 1205-1211 (2021).

19. Mariz, F.C., *et al.* Sustainability of neutralising antibodies induced by bivalent or quadrivalent HPV vaccines and correlation with efficacy: a combined follow-up analysis of data from two randomised, double-blind, multicentre, phase 3 trials. *The Lancet Infectious Diseases* **21**, 1458-1468 (2021).
20. Tsang, S.H., *et al.* Evaluation of serological assays to monitor antibody responses to single-dose HPV vaccines. *Vaccine* **38**, 5997-6006 (2020).
21. Nolan, K.M., *et al.* Development and Validation of Two Optimized Multiplexed Serologic Assays for the 9-Valent Human Papillomavirus Vaccine Types. *mSphere* **8**, e00962-00921 (2023).
22. Kreimer, A.R., *et al.* Efficacy of a bivalent HPV 16/18 vaccine against anal HPV 16/18 infection among young women: a nested analysis within the Costa Rica Vaccine Trial. *Lancet Oncol* **12**, 862-870 (2011).
23. Kreimer, A.R., *et al.* Proof-of-principle evaluation of the efficacy of fewer than three doses of a bivalent HPV16/18 vaccine. *J Natl Cancer Inst* **103**, 1444-1451 (2011).
24. Herrero, R., *et al.* Reduced prevalence of oral human papillomavirus (HPV) 4 years after bivalent HPV vaccination in a randomized clinical trial in Costa Rica. *PLoS One* **8**, e68329 (2013).
25. Safaeian, M., *et al.* Durable antibody responses following one dose of the bivalent human papillomavirus L1 virus-like particle vaccine in the Costa Rica Vaccine Trial. *Cancer Prev Res (Phila)* **6**, 1242-1250 (2013).
26. Hildesheim, A., *et al.* Efficacy of the HPV-16/18 vaccine: final according to protocol results from the blinded phase of the randomized Costa Rica HPV-16/18 vaccine trial. *Vaccine* **32**, 5087-5097 (2014).
27. Lang Kuhs, K.A., *et al.* Reduced prevalence of vulvar HPV16/18 infection among women who received the HPV16/18 bivalent vaccine: a nested analysis within the Costa Rica Vaccine Trial. *J Infect Dis* **210**, 1890-1899 (2014).
28. Lang Kuhs, K.A., *et al.* Effect of different human papillomavirus serological and DNA criteria on vaccine efficacy estimates. *Am J Epidemiol* **180**, 599-607 (2014).
29. Safaeian, M., *et al.* Durability of Protection Afforded by Fewer Doses of the HPV16/18 Vaccine: The CVT Trial. *J Natl Cancer Inst* **110**, 205-212 (2018).
30. Kreimer, A.R., *et al.* Evaluation of Durability of a Single Dose of the Bivalent HPV Vaccine: The CVT Trial. *J Natl Cancer Inst* **112**, 1038-1046 (2020).
31. Tsang, S.H., *et al.* Durability of Cross-Protection by Different Schedules of the Bivalent HPV Vaccine: The CVT Trial. *J Natl Cancer Inst* **112**, 1030-1037 (2020).
32. Shing, J.Z., *et al.* Precancerous cervical lesions caused by non-vaccine-preventable HPV types after vaccination with the bivalent AS04-adjuvanted HPV vaccine: an analysis of the long-term follow-up study from the randomised Costa Rica HPV Vaccine Trial. *Lancet Oncol* **23**, 940-949 (2022).
33. Befano, B., *et al.* Estimating human papillomavirus vaccine efficacy from a single-arm trial: proof-of-principle in the Costa Rica Vaccine Trial. *J Natl Cancer Inst* **115**, 788-795 (2023).
34. Wacholder, S., *et al.* Risk of miscarriage with bivalent vaccine against human papillomavirus (HPV) types 16 and 18: pooled analysis of two randomised controlled trials. *BMJ* **340**, c712 (2010).
35. Kreimer, A.R., *et al.* Efficacy of fewer than three doses of an HPV-16/18 AS04-adjuvanted vaccine: combined analysis of data from the Costa Rica Vaccine and PATRICIA Trials. *Lancet Oncol* **16**, 775-786 (2015).
36. Kreimer, A.R., *et al.* Evidence for single-dose protection by the bivalent HPV vaccine—Review of the Costa Rica HPV vaccine trial and future research studies. *Vaccine* **36**, 4774-4782 (2018).

37. Harari, A., *et al.* Cross-protection of the Bivalent Human Papillomavirus (HPV) Vaccine Against Variants of Genetically Related High-Risk HPV Infections. *The Journal of Infectious Diseases* **213**, 939-947 (2016).
38. Haas, C.B., *et al.* Design and initial findings of a natural history study of anal HPV and associated lesions among young adult women in Costa Rica. *International Journal of Cancer* **157**, 2603-2618 (2025).
39. Porras, C., *et al.* HPV16/18 antibodies 16-years after single dose of bivalent HPV vaccination: Costa Rica HPV vaccine trial. *JNCI Monographs* **2024**, 329-336 (2024).
40. De Carvalho, N., *et al.* Sustained efficacy and immunogenicity of the HPV-16/18 AS04-adjuvanted vaccine up to 7.3 years in young adult women. *Vaccine* **28**, 6247-6255 (2010).
41. Romanowski, B., *et al.* Sustained efficacy and immunogenicity of the human papillomavirus (HPV)-16/18 AS04-adjuvanted vaccine: analysis of a randomised placebo-controlled trial up to 6.4 years. *Lancet* **374**, 1975-1985 (2009).
42. Harper, D.M., *et al.* Efficacy of a bivalent L1 virus-like particle vaccine in prevention of infection with human papillomavirus types 16 and 18 in young women: a randomised controlled trial. *Lancet* **364**, 1757-1765 (2004).
43. Harper, D.M., *et al.* Sustained efficacy up to 4.5 years of a bivalent L1 virus-like particle vaccine against human papillomavirus types 16 and 18: follow-up from a randomised control trial. *Lancet* **367**, 1247-1255 (2006).
44. Roteli-Martins, C.M., *et al.* Sustained immunogenicity and efficacy of the HPV-16/18 AS04-adjuvanted vaccine. *Human Vaccines & Immunotherapeutics* **8**, 390-397 (2012).
45. Zhao, F.H., *et al.* Efficacy, safety, and immunogenicity of an Escherichia coli-produced Human Papillomavirus (16 and 18) L1 virus-like-particle vaccine: end-of-study analysis of a phase 3, double-blind, randomised, controlled trial. *Lancet Infect Dis* **22**, 1756-1768 (2022).
46. Jia, X., Hu, S., Kuang, X. & Qiao, Y. Long-term protective efficacy of the Escherichia coli-produced HPV-16/18 bivalent human papillomavirus vaccine in women vaccinated at 18–45 years: A 9-year follow-up study. *Infectious Medicine* **4**, 100164 (2025).
47. Su, Y., *et al.* Pattern of multiple human papillomavirus infection and type competition: An analysis in healthy Chinese women aged 18–45 years. *Human Vaccines & Immunotherapeutics* **20**, 2334474 (2024).
48. Zhu, F.C., *et al.* Efficacy, immunogenicity and safety of the HPV-16/18 AS04-adjuvanted vaccine in healthy Chinese women aged 18-25 years: results from a randomized controlled trial. *Int J Cancer* **135**, 2612-2622 (2014).
49. Zhu, F., *et al.* Immunogenicity and safety of the HPV-16/18 AS04-adjuvanted vaccine in healthy Chinese girls and women aged 9 to 45 years. *Hum Vaccin Immunother* **10**, 1795-1806 (2014).
50. Zhu, F.C., *et al.* Efficacy, immunogenicity, and safety of the HPV-16/18 AS04-adjuvanted vaccine in Chinese women aged 18-25 years: event-triggered analysis of a randomized controlled trial. *Cancer Med* **6**, 12-25 (2017).
51. Welby, S., Rosillon, D., Feng, Y. & Borys, D. Progression from human papillomavirus (HPV) infection to cervical lesion or clearance in women (18-25 years): Natural history study in the control arm subjects of AS04-HPV-16/18 vaccine efficacy study in China between 2008 and 2016. *Expert Rev Vaccines* **21**, 407-413 (2022).
52. Zhao, F., *et al.* Safety of AS04-HPV-16/18 vaccine in Chinese women aged 26 years and older and long-term protective effect in women vaccinated at age 18-25 years: A 10-year follow-up study. *Asia Pac J Clin Oncol* **19**, 458-467 (2023).
53. Wen, T., *et al.* Ten-year evaluation of HPV vaccine influence on non-vaccine-type-related cervical infections and precancers in Chinese females: follow-up from a randomized control trial. *Journal of the National Cancer Center* **5**, 586-592 (2025).

54. Wen, T., *et al.* The 13-year long-term follow-up on the effectiveness and immunogenicity of the quadrivalent human papillomavirus vaccine in Chinese females vaccinated at 20–45 years of age. *Human Vaccines & Immunotherapeutics* **20**, 2412391 (2024).
55. Zhao, C., *et al.* The eight-year long-term follow-up on the effectiveness of the quadrivalent human papillomavirus vaccine in Chinese women 20-45 years of age. *Hum Vaccin Immunother* **18**, 2052700 (2022).
56. Ault, K.A. & Future II Study Group. Effect of prophylactic human papillomavirus L1 virus-like-particle vaccine on risk of cervical intraepithelial neoplasia grade 2, grade 3, and adenocarcinoma in situ: a combined analysis of four randomised clinical trials. *Lancet* **369**, 1861-1868 (2007).
57. Perez, G., *et al.* Safety, immunogenicity, and efficacy of quadrivalent human papillomavirus (types 6, 11, 16, 18) L1 virus-like-particle vaccine in Latin American women. *Int J Cancer* **122**, 1311-1318 (2008).
58. Garland, S.M., *et al.* Quadrivalent vaccine against human papillomavirus to prevent anogenital diseases. *N Engl J Med* **356**, 1928-1943 (2007).
59. Wheeler, C.M., *et al.* Safety and immunogenicity of co-administered quadrivalent human papillomavirus (HPV)-6/11/16/18 L1 virus-like particle (VLP) and hepatitis B (HBV) vaccines. *Vaccine* **26**, 686-696 (2008).
60. Future II Study Group. Prophylactic efficacy of a quadrivalent human papillomavirus (HPV) vaccine in women with virological evidence of HPV infection. *J Infect Dis* **196**, 1438-1446 (2007).
61. Olsson, S.E., *et al.* Evaluation of quadrivalent HPV 6/11/16/18 vaccine efficacy against cervical and anogenital disease in subjects with serological evidence of prior vaccine type HPV infection. *Hum Vaccin* **5**, 696-704 (2009).
62. Kjaer, S.K., *et al.* A pooled analysis of continued prophylactic efficacy of quadrivalent human papillomavirus (Types 6/11/16/18) vaccine against high-grade cervical and external genital lesions. *Cancer Prev Res (Phila)* **2**, 868-878 (2009).
63. Brown, D.R., *et al.* The impact of quadrivalent human papillomavirus (HPV; types 6, 11, 16, and 18) L1 virus-like particle vaccine on infection and disease due to oncogenic nonvaccine HPV types in generally HPV-naive women aged 16-26 years. *J Infect Dis* **199**, 926-935 (2009).
64. Dillner, J., *et al.* Four year efficacy of prophylactic human papillomavirus quadrivalent vaccine against low grade cervical, vulvar, and vaginal intraepithelial neoplasia and anogenital warts: randomised controlled trial. *BMJ* **341**, c3493 (2010).
65. Munoz, N., *et al.* Impact of human papillomavirus (HPV)-6/11/16/18 vaccine on all HPV-associated genital diseases in young women. *J Natl Cancer Inst* **102**, 325-339 (2010).
66. Haupt, R.M., *et al.* Impact of an HPV6/11/16/18 L1 virus-like particle vaccine on progression to cervical intraepithelial neoplasia in seropositive women with HPV16/18 infection. *Int J Cancer* **129**, 2632-2642 (2011).
67. Joura, E.A., *et al.* Effect of the human papillomavirus (HPV) quadrivalent vaccine in a subgroup of women with cervical and vulvar disease: retrospective pooled analysis of trial data. *BMJ* **344**, e1401 (2012).
68. Future II Study Group. Quadrivalent vaccine against human papillomavirus to prevent high-grade cervical lesions. *N Engl J Med* **356**, 1915-1927 (2007).
69. Kjaer, S.K., *et al.* Final analysis of a 14-year long-term follow-up study of the effectiveness and immunogenicity of the quadrivalent human papillomavirus vaccine in women from four nordic countries. *eClinicalMedicine* **23**(2020).
70. Kjaer, S.K., *et al.* A 12-Year Follow-up on the Long-Term Effectiveness of the Quadrivalent Human Papillomavirus Vaccine in 4 Nordic Countries. *Clinical Infectious Diseases* **66**, 339-345 (2017).

71. Lehtinen, M., *et al.* Head-to-head comparison of two human papillomavirus vaccines for efficacy against cervical intraepithelial neoplasia grade 3 and adenocarcinoma in situ—population-based follow-up of two cluster-randomized trials. *Frontiers in Cellular and Infection Microbiology* **Volume 14 - 2024**(2024).
72. Joura, E., *et al.* Effect of Prior 9-Valent Human Papillomavirus Vaccination on Subsequent Lower Genital Tract Dysplasia After Cervical Excisional Surgery. *Obstetrics & Gynecology* **147**(2026).
73. Castellsague, X., *et al.* End-of-study safety, immunogenicity, and efficacy of quadrivalent HPV (types 6, 11, 16, 18) recombinant vaccine in adult women 24-45 years of age. *Br J Cancer* **105**, 28-37 (2011).
74. Maldonado, I., *et al.* Effectiveness, immunogenicity, and safety of the quadrivalent HPV vaccine in women and men aged 27-45 years. *Hum Vaccin Immunother* **18**, 2078626 (2022).
75. Barnabas, R.V., *et al.* Durability of single-dose HPV vaccination in young Kenyan women: randomized controlled trial 3-year results. *Nature Medicine* **29**, 3224-3232 (2023).
76. Baisley, K., *et al.* Comparing one dose of HPV vaccine in girls aged 9-14 years in Tanzania (DoRIS) with one dose in young women aged 15-20 years in Kenya (KEN SHE): an immunobridging analysis of randomised controlled trials. *The Lancet Global Health* **12**, e491-e499 (2024).
77. Konno, R., Tamura, S., Dobbelaere, K. & Yoshikawa, H. Efficacy of human papillomavirus 16/18 AS04-adjuvanted vaccine in Japanese women aged 20 to 25 years: interim analysis of a phase 2 double-blind, randomized, controlled trial. *Int J Gynecol Cancer* **20**, 404-410 (2010).
78. Konno, R., Tamura, S., Dobbelaere, K. & Yoshikawa, H. Efficacy of human papillomavirus type 16/18 AS04-adjuvanted vaccine in Japanese women aged 20 to 25 years: final analysis of a phase 2 double-blind, randomized controlled trial. *Int J Gynecol Cancer* **20**, 847-855 (2010).
79. Konno, R., *et al.* Efficacy of the human papillomavirus (HPV)-16/18 AS04-adjuvanted vaccine against cervical intraepithelial neoplasia and cervical infection in young Japanese women. *Hum Vaccin Immunother* **10**, 1781-1794 (2014).
80. Yoshikawa, H., Ebihara, K., Tanaka, Y. & Noda, K. Efficacy of quadrivalent human papillomavirus (types 6, 11, 16 and 18) vaccine (GARDASIL) in Japanese women aged 18-26 years. *Cancer Sci* **104**, 465-472 (2013).
81. Koutsky, L.A., *et al.* A controlled trial of a human papillomavirus type 16 vaccine. *N Engl J Med* **347**, 1645-1651 (2002).
82. Mao, C., *et al.* Efficacy of human papillomavirus-16 vaccine to prevent cervical intraepithelial neoplasia: a randomized controlled trial. *Obstet Gynecol* **107**, 18-27 (2006).
83. Rowhani-Rahbar, A., *et al.* Longer term efficacy of a prophylactic monovalent human papillomavirus type 16 vaccine. *Vaccine* **27**, 5612-5619 (2009).
84. Garland, S.M., *et al.* Prior human papillomavirus-16/18 AS04-adjuvanted vaccination prevents recurrent high grade cervical intraepithelial neoplasia after definitive surgical therapy: Post-hoc analysis from a randomized controlled trial. *Int J Cancer* **139**, 2812-2826 (2016).
85. Lehtinen, M., *et al.* Overall efficacy of HPV-16/18 AS04-adjuvanted vaccine against grade 3 or greater cervical intraepithelial neoplasia: 4-year end-of-study analysis of the randomised, double-blind PATRICIA trial. *Lancet Oncol* **13**, 89-99 (2012).
86. Paavonen, J., *et al.* Efficacy of a prophylactic adjuvanted bivalent L1 virus-like-particle vaccine against infection with human papillomavirus types 16 and 18 in young women:

an interim analysis of a phase III double-blind, randomised controlled trial. *Lancet* **369**, 2161-2170 (2007).
87. Paavonen, J., *et al.* Efficacy of human papillomavirus (HPV)-16/18 AS04-adjuvanted vaccine against cervical infection and precancer caused by oncogenic HPV types (PATRICIA): final analysis of a double-blind, randomised study in young women. *Lancet* **374**, 301-314 (2009).
88. Szarewski, A., *et al.* Efficacy of the human papillomavirus (HPV)-16/18 AS04-adjuvanted vaccine in women aged 15-25 years with and without serological evidence of previous exposure to HPV-16/18. *Int J Cancer* **131**, 106-116 (2012).
89. Wheeler, C.M., *et al.* Cross-protective efficacy of HPV-16/18 AS04-adjuvanted vaccine against cervical infection and precancer caused by non-vaccine oncogenic HPV types: 4-year end-of-study analysis of the randomised, double-blind PATRICIA trial. *Lancet Oncol* **13**, 100-110 (2012).
90. Lehtinen, M., *et al.* Ten-year follow-up of human papillomavirus vaccine efficacy against the most stringent cervical neoplasia end-point—registry-based follow-up of *three cohorts from randomized trials*. *BMJ Open* **7**, e015867 (2017).
91. Villa, L.L., *et al.* Prophylactic quadrivalent human papillomavirus (types 6, 11, 16, and 18) L1 virus-like particle vaccine in young women: a randomised double-blind placebo-controlled multicentre phase II efficacy trial. *Lancet Oncol* **6**, 271-278 (2005).
92. Villa, L.L., *et al.* Immunologic responses following administration of a vaccine targeting human papillomavirus Types 6, 11, 16, and 18. *Vaccine* **24**, 5571-5583 (2006).
93. Villa, L.L., *et al.* High sustained efficacy of a prophylactic quadrivalent human papillomavirus types 6/11/16/18 L1 virus-like particle vaccine through 5 years of follow-up. *Br J Cancer* **95**, 1459-1466 (2006).
94. Skinner, S.R., *et al.* Efficacy, safety, and immunogenicity of the human papillomavirus 16/18 AS04-adjuvanted vaccine in women older than 25 years: 4-year interim follow-up of the phase 3, double-blind, randomised controlled VIVIANE study. *Lancet* **384**, 2213-2227 (2014).
95. Wheeler, C.M., *et al.* Efficacy, safety, and immunogenicity of the human papillomavirus 16/18 AS04-adjuvanted vaccine in women older than 25 years: 7-year follow-up of the phase 3, double-blind, randomised controlled VIVIANE study. *Lancet Infect Dis* **16**, 1154-1168 (2016).
96. Giuliano, A.R., *et al.* Efficacy of quadrivalent HPV vaccine against HPV Infection and disease in males. *N Engl J Med* **364**, 401-411 (2011).
97. Goldstone, S.E., *et al.* Efficacy, immunogenicity, and safety of a quadrivalent HPV vaccine in men: results of an open-label, long-term extension of a randomised, placebo-controlled, phase 3 trial. *Lancet Infect Dis* **22**, 413-425 (2022).
98. Hillman, R.J., *et al.* Immunogenicity of the quadrivalent human papillomavirus (type 6/11/16/18) vaccine in males 16 to 26 years old. *Clin Vaccine Immunol* **19**, 261-267 (2012).
99. Palefsky, J.M., *et al.* HPV vaccine against anal HPV infection and anal intraepithelial neoplasia. *N Engl J Med* **365**, 1576-1585 (2011).
100. Palefsky, J.M., *et al.* Anal Human Papillomavirus Infection Progression to Disease Among Men Who Have Sex With Men. *Clinical Infectious Diseases* **82**, e616-e625 (2026).
101. Giuliano, A.R., *et al.* Design of a multicenter, randomized, double-blinded, placebo-controlled phase III trial evaluating the 9-valent human papillomavirus (HPV) vaccine to prevent persistent oral HPV infection in men living with human immunodeficiency virus: ULACNet trial 201. *Vaccine* **61**, 127447 (2025).

102. Safaeian, M., *et al*. Direct Comparison of HPV16 Serological Assays Used to Define HPV-Naïve Women in HPV Vaccine Trials. *Cancer Epidemiology, Biomarkers & Prevention* **21**, 1547-1554 (2012).
